# Subtyping Depression using Brain-Gut Electrophysiology for Early Prediction of Antidepressant Response: a multicentric clinical study

**DOI:** 10.1101/2025.03.07.25323496

**Authors:** Amal Jude Ashwin Francis, Alok Bajpai, Nandini Priyanka Balasubramani, Hari Prakash Tiwari, Shikha Singh, Kritika Chawla, Ayushi Devendra Singh, Dhananjay Chaudhari, Pragathi Priyadharsini Balasubramani

## Abstract

Depression affects approximately 5% of adults worldwide, with India reporting a prevalence of 4.5%. Oral medication is a common treatment, but over 50% of patients fail to respond to the first-line antidepressants and often require medication adjustments or augmentation. This highlights the urgent need for predictive models that can guide personalized treatment strategies more quickly. Our study aims to achieve three key objectives: first, to assess the predictive ability of previously identified biomarkers such as frontal theta power and alpha asymmetry, in explaining the response to interventions within our sample; second, to evaluate the utility of whole-person research approach, focusing the gut-brain interactions, in predicting responses; and third, to identify reliable early biomarkers that can predict responses across various phenotypic subtypes. Across two sites, a total of 161 (+45) participants, including 99 (45) treatment-naive patients, enrolled in our study from site 1 (+site 2) which spanned three visits. We aimed to predict antidepressant outcomes at the third visit (4-6 weeks) using data collected from visits one (baseline) and two (7-10 days), and the data from site 2 was solely used for testing the predictive utility.

Our predictive models, which incorporated electrophysiological data from both the brain and gut along with clinical information, achieved an cross validation (independent test) performance of 78% (80%) specificity and 84% (71.43%) sensitivity in identifying non-responders to antidepressant treatment administered as per Clinical Practice Guidelines of India. We found that certain electrophysiological features were strongly predictive of treatment outcomes for specific depression subtypes. For example, increased excitation-inhibition ratios in the fronto-central brain regions were predictive for patients with dominant anxiety and sleep symptoms. Similarly, decreased tachygastric gut coupling with the sensory-motor brain region predicted treatment non-response in patients with high levels of negative self-thoughts. Increased connectivity in the right fronto-central region was associated with better outcomes in patients with significant appetite issues. Additionally, higher fronto-central theta power and beta asymmetry were predictive of responses in patients with a composite set of symptoms.

Our findings suggest that combining brain and gut electrophysiological markers with clinical phenotyping offers a promising, scalable approach to personalize depression treatment. This approach could guide clinicians in developing more effective and tailored medication strategies, ultimately improving patient outcomes.

## 1. INTRODUCTION

According to the World Health Organization (WHO), Major Depressive Disorder (MDD), commonly known as depression, is a mental health condition characterized by persistent feelings of sadness, lack of motivation, and an inability to experience pleasure. Key symptoms of depression include disrupted sleep and appetite, difficulty concentrating, feelings of hopelessness, low energy, and thoughts of suicide. The current standard treatment for depression involves selective serotonin and norepinephrine reuptake inhibitors (SSRIs/SNRIs). However, this approach typically follows a trial-and-error process, where patients are prescribed medications, but it can take 4 to 6 weeks to assess their effectiveness. Unfortunately, non-response or non-remission rates are high—around 50-60%—following the initial treatment trial. Many patients either experience side effects or find that the medication doesn’t effectively alleviate their symptoms, which leads to further adjustments or changes in the treatment regimen (Bauer et al., 2007; Fochtmann & Gelenberg, 2005; Pigott et al., 2010; Quitkin et al., 2003).

There is a notable gap in the availability of neurophysiological tools that can accurately predict treatment outcomes within a short timeframe—less than two weeks—while accounting for the neuroplastic effects of depression. Earlier attempts utilize a combination of clinical health records, behavioral data, and genomic information to achieve a commendable efficiency rate of around 72% within a 4-week timeframe (Athreya et al., 2021; Bares et al., 2015; Jaworska et al., 2019; John Rush et al., 2006; Keitner et al., 2008; Leuchter et al., 2009; Trivedi et al., 2006; Warden et al., 2007). Relatedly, there are a few studies that look into neurophysiology at any timepoint to predict depression severity (Cao et al., 2019; Hasanzadeh et al., 2019; Sadat Shahabi et al., 2021), and claim to achieve greater than 90% accuracy.

Large randomized clinical trials, such as EMBARC have shown that higher baseline theta-band activity in the anterior cingulate cortex and its connectivity with salience-related brain networks are associated with better responses to sertraline treatment (Whitton et al., 2019). Data from the STAR*D study further emphasize the importance of early response as a predictor of remission with citalopram treatment (Kubo et al., 2023). Several studies, nearly a decade old, tracked changes in neural activation due to medication and found that increased frontal theta power and reduced frontal alpha asymmetry were key markers of treatment response, even for medication-resistant patients, achieving over 91% accuracy when combined with depression rating scale features (Bares et al., 2015). Moreover, frontal beta power has also been linked to depression (Knott et al., 2001; Lieber & Prichep, 1988).

However, more recent studies over the past five years have presented contradictory findings. Some research suggests that frontal alpha asymmetry is not sensitive to treatment response (Kołodziej et al., 2021; Lee et al., 2020; van der Vinne et al., 2019), while other studies argue that it’s not low-frequency power increases, but rather aperiodic activity, that better explains treatment responses in patients (Smith et al., 2023). These conflicting views raise concerns about the reliability and robustness of previously identified neurophysiological markers. Several clinicians have called for a reform in depression treatment strategies, incorporating biological insights, as outlined in the Research Domain Criteria (Kas et al., 2025). A more comprehensive phenotypic profiling of neural circuits and physiological mechanisms, with precision intervention targets, may significantly enhance remission rates in depression treatment.

In recent years, growing evidence suggests that depression severity is not solely reflected in brain physiology but also in whole-body physiology. Gut dysfunctions, such as irritable bowel syndrome and gastroparesis, often confound depression symptoms (Lydiard, 2001; Masand et al., 1995; Overs et al., 2024; Zamani et al., 2019). This has led to an increasing recognition of the gut-brain interaction as crucial to understanding depression symptoms (Drossman, 2016; Goyal et al., 2021). Despite this, gut-related markers have not been incorporated into depression research or clinical treatment strategies.

Our study introduces a novel approach by integrating whole-body cognition, including gut-brain coupling and electroencephalography (EEG), to predict treatment response. Additionally, we incorporate cognitive task probes to better understand how neural circuits are recruited during various tasks, which can aid in predicting treatment outcomes. Specifically, we hypothesize that combining physiological data from the brain and gut during cognitive tasks, along with clinical and demographic information, can enable early prediction of treatment outcomes in depression— well before the typical 4-6 week assessment period. A key aspect of our work is the identification of phenotypic subtypes within depression, which has allowed us to uncover distinct neurophysiological markers linked to specific symptom clusters. We found that different subtypes of depression, defined by symptom dominance (e.g., anxiety, sleep issues, negative self-thoughts, or appetite changes), show unique brain and gut signatures that predict treatment response.

By combining phenotypic profiling with electrophysiological biomarkers, our predictive model can offer personalized treatment guidance—identifying early responders and non-responders as early as 7-10 days into treatment. This contrasts with the traditional 4-6 week evaluation period and could significantly enhance clinicians’ ability to tailor interventions more effectively and quickly. Ultimately, our study offers a scalable, precision-based framework that integrates both brain and gut data, as well as clinical phenotyping, to improve treatment outcomes in depression.

## 2. METHODOLOGY

The methodology section can be broadly divided into 4 sub-sections. 1) Data collection: This sub-section covers the experimental setup and tools used to collect longitudinal electrophysiological data and the self-reported questionnaires. 2) Feature extraction: Using the collected electrophysiological data, the quantitative features extracted are discussed elaborately in this sub-section. 3) Feature selection: The approaches used to select the relevant features for model development and analysis are defined in this sub-section. 4) Model development: Finally, the Machine Learning models developed for classifying clinical participants into responders/non-responders are outlined in the last sub-section of the methodology section.

### 2.1. Data Sites, Ethical clearance

The clinical study data was acquired from two independent sites. We developed our model prototype with the data acquired at Indian Institute of Technology, Kanpur site (Site 1), in collaboration with the institute psychiatrist who sees patients via. his psychiatry clinic. Ethical clearance for the study was obtained by the Institutional Ethics Committee of Indian Institute of Technology Kanpur in February 2022.

Based on the above study, a clinical trial was planned at an independent site, GSVM Medical College, Kanpur (Site 2) and registered in Clinical Trial Registry of India (CTRI/2025/04/084404). Ethical approval from the GSVM ethics committee was obtained under the registration number EC/BMHR/2025/28. The data acquired from this site were used solely for an entirely held-out evaluation of the previously developed predictive model presented in this manuscript.

The informed consent from the participants were obtained orally and in a written format. The participants were awarded coupons for each visit and were allowed to voluntarily quit the study at any time without any deduction in compensation for the visit. The data confidentiality was maintained and a subject ID was assigned to the participants to mask their what and whereabouts before further processing of data.

### 2.2. Inclusion criteria

Healthy young adults from the university campus and community settings such as schools and colleges were included as a control group. Treatment naive patients were recruited for depression group. Participants were included from all Handedness and Genders. Depressed participants received unimodal medication treatment during the time of study.

### 2.3. Exclusion criteria

Patients who are in their Pregnancy or Postpartum period, with any neurological or medical comorbidity that could influence research investigations were excluded from the study. Additionally for depressed patients, participants with suicidal intent or any psychiatric emergency, bipolar depression, psychotic symptoms and substance use disorder were excluded from the study. Participants with multimodal treatment strategy were excluded from the study.

### 2.4. Participants consort

From the site 1, out of 161 people who participated in the study, electrophysiological data of brain (electroencephalogram) and gut (electrogastrogram) were collected from 138 participants including both control and patient population at the baseline visit. However, only 62 participants turned up for all three visits and EEG and EGG were collected from them, where visit 1 (baseline visit) was at the day 0 of participant visit, visit 2 (intermediate follow-up visit) between 7-14 days from then, and the next visit 3 (final follow-up visit) after 30-40 days (refer to **Figure 1A**). Control population corresponds to the population that is not seeking any clinical help and are not diagnosed with any mental disorders.

**Figure 1:**
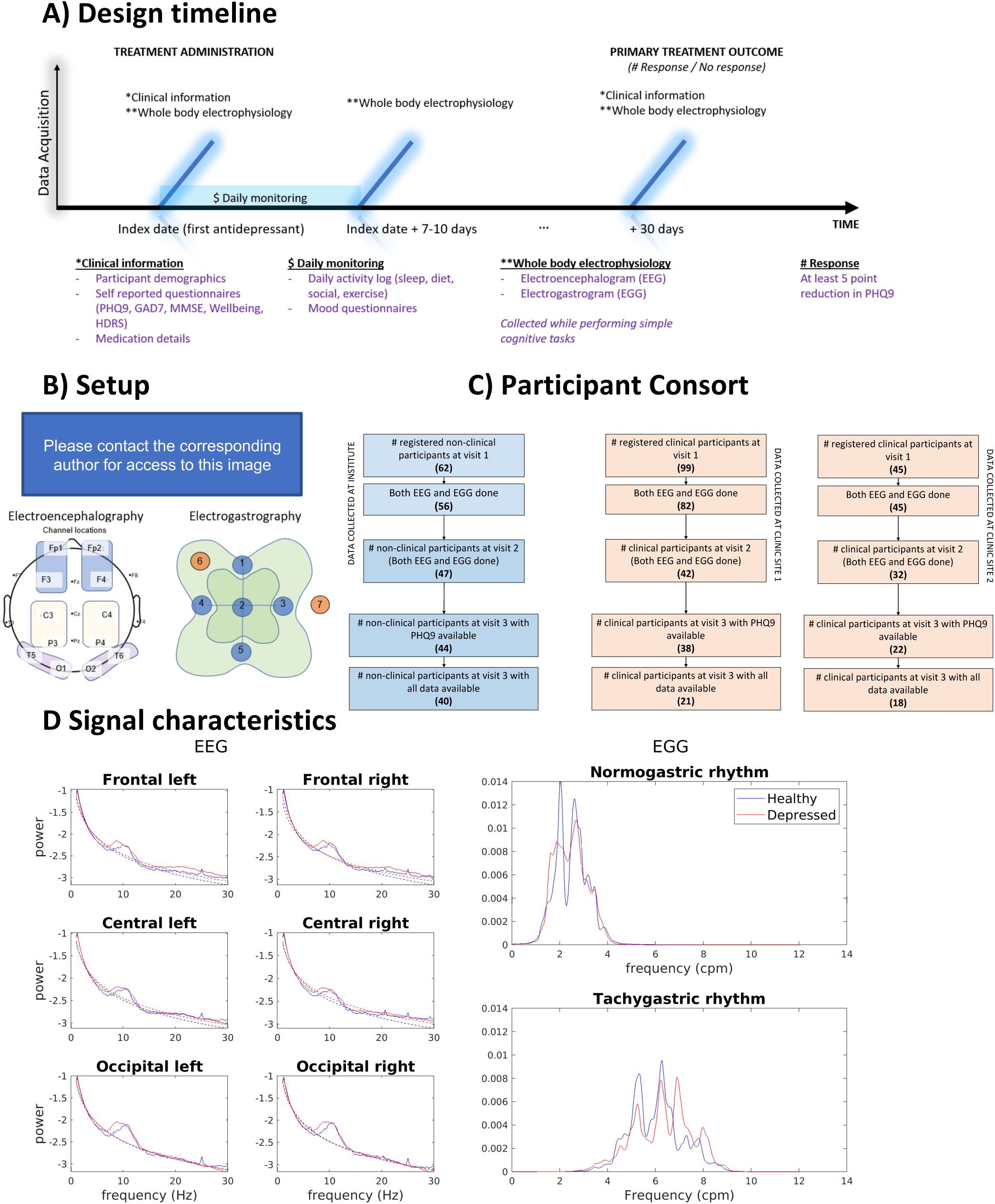
Study design and baseline electrophysiology characteristics. **A)** Study timeline: In visit 1, clinical information such as demographics, questionnaires and baseline whole body electrophysiology (electroencephalography-EEG, electrogastrography-EGG) data are collected. During visit 2, only whole-body electrophysiology while performing simple cognitive tasks is collected. In the final visit, self-reported questionnaires and whole-body physiology are collected. **B)** Experimental setup: The grouping of EEG electrodes in 6 regions and the location of electrodes for electrogastrogram set-up are depicted. **C)** Participant consort: The number of participants for each visit with electrophysiology and PHQ9 questionnaire data are highlighted. **D)** Averaged power spectral density curves for clean EEG and EGG across tasks: Log transformed PSD and aperiodic fit (dotted lines) for the filtered band range of healthy and depressed population are plotted for comparison, central theta EEG were significantly greater in the depressed population. In EGG, we present the normogastric (0.03-0.07Hz) and tachygastric (0.07-0.15Hz) power for the two groups, and the normogastric signals were higher in control.

From the site 2, 45 patients registered totally by October 2025, 32 completed visit 2, 22 completed the self-reports in visit 3, in which only 18 of them provided their physiology data (EEG, EGG) in visit 3.

Patient population corresponds to the participants who are clinically diagnosed to have depression, naïve in medicine intake, and are intaking clinical assistance specifically through oral medication (refer to **Figure 1C**). In case of antidepressant history in patients, careful medication strategies such as tapering with 1 month washout period was used to tailor to individual medication history of the patients. The medication was chosen according to the judgement of the attending physician and in accordance with the Clinical Practice Guidelines (CPG, Gautam et al, 2017). All the participants had normal eye-sight or corrected to vision eye-sight. No other treatments were prescribed to the patient population or control population. A small token of gratitude was provided to the control group for their time. As the study was conducted in a real-world setting, choice of medications were not controlled by the experimenters.

### 2.5. Cognitive tasks setup

The participants performed simple tasks that have been reported to evoke various cognitive processes such as interoception, attention, memory, arithmetic calculation and language. Participants were asked to perform certain tasks on all 3 visits, while the other tasks were performed only on the second visit. Resting state eyes open and eyes closed tasks were performed for approximately 5 minutes each while resting on the chair in a quite setup. An interoceptive breathing task was then performed where the participants were asked to focus on their breathing, hyperventilate and deep breath by holding it for about 5 secs for approximately ten times spanning a total of 2 to 3 minutes. Varying frequency photic task was administered, where the photic stimulus that is present at the right top location of the participant flickers at the rate of 3,5,7,9,11,13,15,17,19,21 Hz for 10 seconds each frequency, inter-frequency-interval of 1 sec, and a total of 110 secs. The above mentioned 4 tasks were performed on all the 3 visits. During visit 2 and 3, in addition to the 4 tasks, the participants performed picture description task, Counting backwards in multiples of 7 from 100 to 40 in an arithmetic task. Mini mental state examination was performed by the participant and the results of the same were stored to assess different cognitive abilities of the participants.

### 2.6. Data acquisition

We administered self-report questionnaires such as PHQ9 (Kroenke et al., 2001), GAD7 (Spitzer et al., 2006), MMSE (Arevalo-Rodriguez et al., 2015), expert administered HDRS (Hamilton, 1960), during visits 1 and 3, wellbeing (Tennant et al., 2007), along with collection of electrophysiological measures. A daily activity log was administered between visits 1 and 2, whose data was not used in the current study. Electrophysiology signals were obtained while the participant performed all the simple cognitive tasks resting comfortably in a chair. Electroencephalogram (EEG) was recorded using the 24-channel montage with 22 recording electrodes, 1 reference, 1 ground electrode following 10-20 system from manufactured by Clarity medicals in the name of BrainTech machine. The sampling rate of the data collected was 256 Hz. The Electrogastrogram (EGG) was recorded using an OpenBCI setup with 1 ground, 1 reference, 2 recording electrodes, placed as mentioned in the figure (refer to **Figure 1B**). The data was acquired at a sampling rate of 250Hz. The same setup was used for all three visits. For about 23 subjects who were recruited during the final stages of the study, a 7 channel EGG setup was used, while for the remaining subjects, a 5-channel setup was used.

### 2.7. Electrophysiology data preprocessing

#### 2.7.1 EEG

EEG data cleaning was performed using EEGLAB v2022.1, MATLAB R2022b.

### Bandpass filtering

The EEG data was bandpass filtered using a Hamming windowed FIR filter (pop_eegfiltnew function in EEGLAB) with ‘locutoff’, ‘highcutoff’ and ‘filtorder’ parameters set to 0.5,35 and 3300 respectively. Slow frequency drifts and high frequency channel noise are removed during bandpass filtering.

### Labeling bad channels

Bad channels were labeled post filtering based on whether the standard deviation of any channel data exceeds the 75^th^ percentile of standard deviation of the rest of the channel and is greater than 100 or less than 1 microVolts.

### Source localization-based artifact removal

Independent Component Analysis (ICA) was executed for the data without the bad channels using pop_runica function with ‘icatype’ parameter set as runica. For *n* number of channels, ICA returns at most *n* number of components. The labels for the components were extracted using iclabel function in EEGLAB which classifies the components into **‘brain’, ‘eye’, ’muscle’, ‘heart’, ‘line noise’, ‘channel noise’, ‘others’** and returns the predictive probabilities of the components for each class. Bad components, the ones which have sum of predictive probability of the brain and others to be less than 0.1, were removed from the channel signals using pop_subcomp function in EEGLAB.

### Interpolation of bad channels

The labeled bad channels were interpolated using pop_interp function with the ‘method’ parameter set to spherical.

### Common average referencing

Finally, common average re-referencing was performed using pop_reref function. The common noise that is recorded by all the channels is reduced due to the common average referencing method.

### Z-score normalization

The resulting EEG signals for a task and a subject was Z-score normalised across all channels.

#### 2.7.2. EGG

#### Bandpass filtering

To obtain normogastric signal, the EGG signal was bandpass filtered between 0.03 (1.8cpm) and 0.07 (4.2cpm) using the fir2 function in MATLAB with a transition width of 0.01 and filter order of 3. For tachygastric signals, the lower and upper frequencies were set to 0.07 (4.2cpm) and 0.15 (9cpm) respectively (Wolpert et al., 2020). The power spectral densities of the filtered signals are used to validate the process of filtering (refer to **Figure 1D**).

### 2.8. Quantitative electrophysiology feature extraction

The data was fragmented into 1minute fragments and quantitative features were extracted using preprocessed EEG and EGG. The EEG features were grouped region-wise into 6 regions, frontal right (Fp2, F4), frontal left (Fp1, F3), central right (C4, P4), central left (C3, P3), occipital right (O2, T6) and occipital left (O1, T5).

#### 2.8.1. Absolute band power of brain signals

The power spectral density was computed by considering the Z-score normalized channel signal and performing fast fourier transform with window size as 5 seconds and 50% overlap. The average band power (theta (4-7 Hz), alpha (8-12 Hz) and beta (13-30 Hz)) were calculated within the range as the absolute band power.

#### 2.8.2. Relative band power of brain signals

The relative band power was computed by dividing the absolute band power and total power between 1 to 35Hz because it encompasses the bands considered and is also within the bandpass filtering range.

#### 2.8.3. Separating the aperiodic and Periodic band power of brain signals

The Power spectral density (PSD) was obtained using pwelch function in MATLAB considering 5 second windows and 50% overlap between windows and were normalized using eqn. (1). Brain signals that are recorded using EEG has a 1/f component i.e., there is more power at lower frequency and less power at higher frequency. Therefore, fitting oscillations one over frequency (FOOOF) method was used to separate the aperiodic and the periodic components in the power spectral density (Donoghue et al., 2020). FOOOF fits 2 aperiodic parameters, exponent and offset, to the log transformed power spectral density of the broadband EEG signal and generates an aperiodic component. Removing the aperiodic component, the periodic component is obtained that is comparable across frequencies. Band powers were computed using only the periodic component. The aperiodic parameters, offset and exponent, were also stored after fitting oscillations one-over frequency.

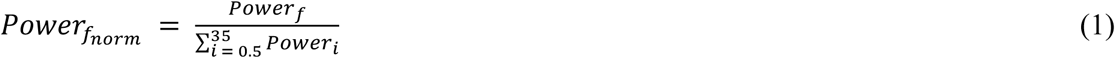

#### 2.8.4. Approximate entropy

Approximate entropy is a measure of randomness in the signal. It was calculated using the approximateEntropy function in MATLAB and the lag was set to be 1 second. A higher value of approximate entropy means higher randomness and a lower value means higher predictability and less randomness.

#### 2.8.5. Theta cordance

Historically, theta cordance has been used as a measure of energy consumption in a given region. It is calculated by taking the average of absolute theta power and relative theta power as computed in sections 2.8.1 and 2.8.2 respectively.

#### 2.8.6. Band power asymmetry

Band power asymmetry between right and left region is the difference of their relative band powers. A negative value indicates that the band power is greater in left region and a positive value indicates that the band power is higher in the right part of the region.

#### 2.8.7. Magnitude squared coherence - a measure of functional connectivity

Magnitude squared coherence computes the cross power spectral density between 2 signals and provides insights about the similarity between the PSD of 2 signals. If a particular frequency is present in signal A and signal B, the coherence at that particular frequency is closer to 1 and similarly if it is absent in both, it is closer to 1. In any other case, the coherence value is low and tends towards 0. In our study, coherence was computed using the mscohere function in MATLAB and using the clean broadband EEG signals of the electrodes. The coherence at specific bands were computed by taking the average coherence at the frequencies of interest.

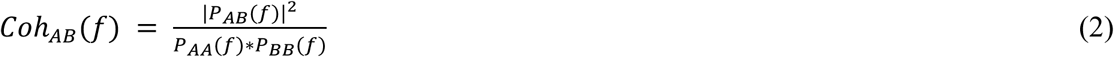

where the numerator term is the cross power spectral density between signal A and B at frequency ‘f’ and the denominator is the normalizing term used to limit the coherence value from 0 to 1.

#### 2.8.8. Weighted - phase lag index

Weighted PLI between 2 signals is a measure of presence of consistence phase difference between after accounting for the effect of noise and volume conduction to an extent (Vinck et al., 2011). If there is a consistent phase difference between 2 signals, commonly referred to as phase synchronization, then the value of wPLI tends to 1, otherwise, it will tend towards 0.

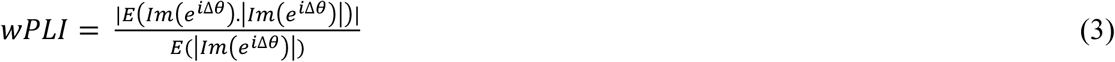

where Δ𝜃 is the instantaneous phase difference computed by using the hilbert transform of the signals and 𝐼𝑚I𝑒^RΔS^N is the imaginary part of the complex phase difference.

#### 2.8.9. Band power of gut signals

The peak band power for the filtered gut signal was computed using the pwelch function in MATLAB with 1 minute window and 50% overlap. The peak frequency corresponds to the frequency at which the peak power is observed.

#### 2.8.10. Phase amplitude coupling between EEG and EGG - a measure of gut-brain coupling

The collected EEG and EGG signals were phase locked at the level of seconds and the analytical signal was constructed using the instantaneous amplitude of the higher frequency EEG signal and the instantaneous phase of the lower frequency EGG signal. The instantaneous amplitude was computed using the absolute of the hilbert transform of the EEG signal and the instantaneous phase was computed using the angle of the hilbert transform of the Z-score normalized, filtered EGG signal. Phase amplitude coupling (PAC) for the analytical signal was computed using the following formula,

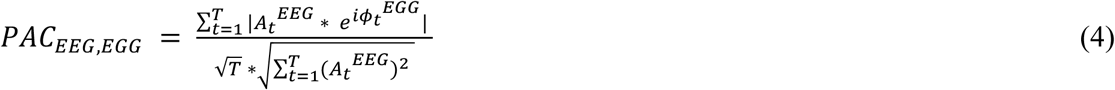

The PAC measure was computed using the timepoints belonging to the top 5 percentile of instantaneous amplitude to represent gut-brain coupling.

### 2.9. Feature scrutinization for Prediction model - criteria based

The extracted features were tested for inter-feature correlation. Highly correlated features (>0.8) were merged by taking their average and the final set of features were passed for criteria-based scrutinization as mentioned in the following section. Even datapoints of participants with missing visits were included for this process. These feature groups values closely represent the top principal component with Pearson *corr > 0.98*.

*Criteria (i): Does the feature show differences between healthy and depressed individuals?* The extracted electrophysiology features from the baseline visit recordings were tested for statistical difference between healthy and depressed populations based on mental health scores (PHQ9, healthy: baseline PHQ9 <=5, depressed: baseline PHQ9 > 5). If the p-value was less than 0.05, then the feature passes the criteria.

*Criteria (iia): Do the change in feature value over time reflect mental health severity?* The change of the electrophysiological feature values at the intermediate follow-up visit from baseline (visit 2 - visit 1) were correlated with the change in PHQ9 scores at final follow-up visit from baseline (visit 1 - visit 3). If the correlation coefficient was significant (p-value<0.05), then the feature qualifies the criteria.

*Criteria (iib):* In the same lines as before, the change in any feature value at the final follow-up visit from baseline (visit 3 - visit 1) were correlated with the change in PHQ9 score at final follow-up visit from baseline (visit 1 - visit 3). If the correlation coefficient was significant (p-value<0.05), then the feature is said to qualify the criteria.

The features that satisfy one of the above criteria were filtered for the predictive modeling. For baseline features, if criteria (i) is satisfied, the baseline electrophysiological feature was used for model development. On feature plasticity, if criteria (iia) or criteria (iib) is satisfied, then the change in feature value at intermediate follow-up visit from baseline visit (visit 2 - visit 1) is considered.

The final set of features used for predictive model development are presented in **Supplementary table A**.

### 2.10. Feature generation for the prediction models

The criteria-based scrutinized quantitative electrophysiology features, along with the demographics of the participants and the individual answers to the questionnaires such as PHQ9, GAD7 and MMSE recorded at visit 1 and visit 2 were used as input to develop the predictive models. For the electrophysiology data fragmented into 1-minute fragments, corresponding demographics and scores of each subject were repeated for all the fragments. The fragments of various tasks were concatenated such that the 1st minute of a particular task was concatenated with the 1st minute of another task.

*Feature Imputation:* For feature categories such as baseline EEG, baseline EGG, change in EEG and change in EGG with missing data for less than 2 tasks and shorter tasks, the missing values were imputed using other features of the same category using Multiple Imputation using Chained Equations algorithm. IterativeImputer function from sci-kit library in PYTHON software, where the values were iteratively imputed in a round-robin fashion for a maximum of 10 iterations until the values reach an asymptotic stability. Features were generated for 5 fragments in any task, the average inter-fragment correlation across features and groups was 0.77±0.19.

### 2.11. Stepwise testing of feature categories for predicting treatment response

In order to illustrate the significant incremental variance explained by the longitudinal and gut-brain coupling features, in addition to the classical baseline EEG, we sequentially added selected longitudinal EEG and gut-brain coupling (PAC) feature groups and predicted the change in PHQ9 score at final follow-up visit from baseline (visit3-visit1) using Ridge Regression. As baseline EEG and scores are easy to obtain in a clinical setting, they were tested initially. Due to abundance of studies and accessibility to EEG in the clinics, longitudinal EEG was added next. Finally, baseline and longitudinal gut features were added to the input features. For each category, Principal Component Analysis was performed to reduce the dimensions and the components required to explain 70% data variance were used as input to RidgeRegression. The performance of the model was assessed using leave one subject out cross validation strategy.

### 2.12. Model outcome operationalization

Five models were developed for comparing the performance and explainability of treatment outcome prediction. The model outcomes were Patients with significant change (SC) or no change (NC) or healthy controls: That is, if there was >=30% reduction in final follow-up PHQ- 9 score as compared to the baseline visit or if the final follow-up (visit3) PHQ9 was <=5, then there is positive change in mental health and the subject was considered as a “Significant Change” (SC); the 30% reduction as model response criteria assisted to maintain class balance for training and testing the machine learning models, and also accounted for early assessment of change in just a week’s time. This is in contrast to the standard clinical routine of assessing response of >50% in about 4-6 weeks timeline from the start of the treatment. If not, then the output is treated as no change in mental health and the subject was considered as a “No Change” (NC). Healthy controls are non-clinical participants with baseline PHQ9 <=5.

Models 1 and 2 are Multi-Layer Perceptron that uses all the scrutinized electrophysiological features along with demographics and questionnaires, and outputs whether the subject is a no change in mental health/significant change in mental health (Model 1) and NC/SC/HC (Model 2) based on baseline PHQ9 score and change in PHQ9 scores at the baseline visit and final follow-up visit. The MLP was built using 2 hidden nodes with tanh activation function with constant learning rate of alpha=0.001. The parameters were finalized using Grid search algorithm.

Model 3 is a logistic regression model that outputs whether a depressed subject is a SC/NC. For the development of this model, only the data points of subjects whose PHQ9 score at baseline is >5 was used. Therefore, all the healthy controls were excluded and only 2 classes were used in this model.

Models 4 and 5 are Recurrent Neural Networks that learns the time dynamics signature differentiating various subject groups, with input as the 1 minute fragments through time and had a design of 2 hidden nodes to the output layer with either 2 nodes representing NC, SC or 3 nodes representing NC, SC, HC, respectively.

Oversampling and Synthetic Minority Oversampling Technique (SMOTE) was used by all the models to address the issue of class imbalance and increase the datapoints in the training dataset respectively. The imblearn library in PYTHON was used with the default parameters of sampling_strategy as “auto” and n_neighbours as 5. Only the minority classes were oversampled to match the number of samples in the majority class.

### 2.13. Baseline questionnaire based subtyping of phenotypes using unsupervised Kmeans

The distribution of baseline questionnaires (PHQ9, GAD7, MMSE) was presented in a reduced dimensional form using Kmeans clustering algorithm with number of clusters set as 3 (optimal clusters identified using elbow and silhouette methods). Each cluster was labelled as HC, SC or NC if their data samples form 2/3^rd^ majority in the cluster.

### 2.14. Supervised Subtyping based on symptoms

Further, we cluster the phenotype presentations in our participants into gut symptoms or homeostatic regulation issue dominant symptom presentation, cognitive behavioral issues dominant phenotype, and arousal related presentations. Specifically, we categorize any patient as symptom dominant if their symptom score is greater than the population median.

Sleep score was computed as the mean of PhQ questions 3 (*Trouble falling or staying asleep, or sleeping too much*) and 4 (*Feeling tired or having little energy*), appetite as PhQ q5 (*poor appetite of overeating*), negative thoughts about self as PhQ q6 (*that you are a failure or have let yourself or your family down*), social symptoms as the average of questions 7 (*Trouble concentrating on external things*) and 8 (*Moving or speaking so slowly that other people could have noticed*), and anxiety as the average score in GAD7.

### 2.15. Feature importance

SHAPley value is a measure of marginal contribution of the feature towards the prediction of a class for a given datapoint. Depending on the feature value, the SHAPley value increases or decreases, and this relationship is captured by the slope of the linear model fit between the feature value of the all the datapoints and their corresponding SHAPley values. The feature importance score is calculated using the following formula,

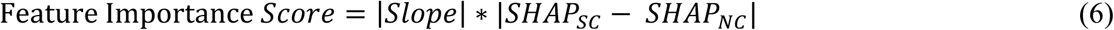

In the above formula, the score is maximum when the magnitude of slope is maximum and marginal contribution of the feature increases the prediction of one class (SC or NC) while bringing down the prediction of the other.

### 2.15. Statistics

For all correlation analysis, the test for normality was performed using Anderson-Darling test and if the data was normal, the correlation was calculated using Pearson correlation method. If the feature failed to pass Anderson-Darling test, then Spearman rank-based correlation method was used. As demographics have categorical variables, chi-square test was performed to compute statistical significance.

For feature comparisons based on existing theories, based on normality, t-test or rank-sum test was performed to calculate the p-value. As the validation of existing theory was not a part of our hypothesis testing, no corrections were performed to the p-values.

## 3. RESULTS

Our primary study objective is to assess whether any behavioral or whole person neurophysiology markers can predict the treatment outcome as soon as 7-10 days after intake of medicine in depression patients? To answer, we setup our experiment in a longitudinal design and recruited treatment naïve patients, control subjects; the patients start their medicine intake from the index date on visit 1 to the clinic, follows up in 7-10 days time for visit 2 experimental procedures, and again after 30-40 days for follow up visit 3. These patients were prescribed to take Selective Serotonin Reuptake Inhibitors (SSRI), Benzodiazepines or atypical antidepressants. The age range-matched controls were recruited from the community outside of the clinic and were assessed in our research laboratory setup. **Figure 1** presents the schematic of the experimental timeline, the setup and the consort diagram. We collected initial patient information, demographics, medicine information, history of trauma, during the index visit 1 date. During visits 1 and 3, we collected electrophysiological data including electroencephalogram (EEG), electrogastrogram (EGG) for various cognitive tasks, along with administration of PHQ9, GAD7, MMSE, and conducted HDRS clinical interview. During visit 2, we collected their EEG, EGG for cognitive tasks, and collected wellbeing scores. The cognitive paradigms include being in simple eyes open resting state, eyes closed resting, performing hyperventilating interoception, and photic stimulation, that were administered to all participants in all 3 visits.

All the initial data analysis and model development presented in this manuscript were based of the data collected from site 1. The control population had a mean age of 34.3 yrs (±12.17) with 51 male participants and 10 female participants. The patient population has a mean of 35.4 yrs (±15) with 73 male participants and 26 female participants. The signal characteristics **(Figure 1D)** shows that broadly across visits, there are no statistically different spectral EEG presentation between control and depressed population when averaged across regions, however, theta power in central regions were significantly greater for the depressed population when compared to the control population (p=0.014 (left), 0.045 (right). The control population had a higher normogastric EGG power (p=0.048, **Table 1).**

**Table 1:** Terminologies used to define the participants based on their symptomological profile. The groups are categorized as HC/SC/NC and are used for model development and analysis. The rows indicate distinct sites of participant recruitment, while the first column indicates the status of initial visit 1 PHQ9, and the second and third columns indicate the final visit 3 PHQ9 scores.

| Site of Recruitment | PHQ9≤5 (visit 1) | PHQ9$>$5 (visit 3), ≥30% reduction (visit 1-3) | PHQ9$>$5 (visit 3), $<$30% reduction (visit 1-3) |
| --- | --- | --- | --- |
| Non-clinic (control) | Psychotypical healthy controls (HC) | Non-clinical responders | Symptomatic controls (NC) |
| Clinic (patient) | Sub-threshold Depression | Significant change in Mental Health (SC) | No change in Mental Health (NC) |

**Table 2:**
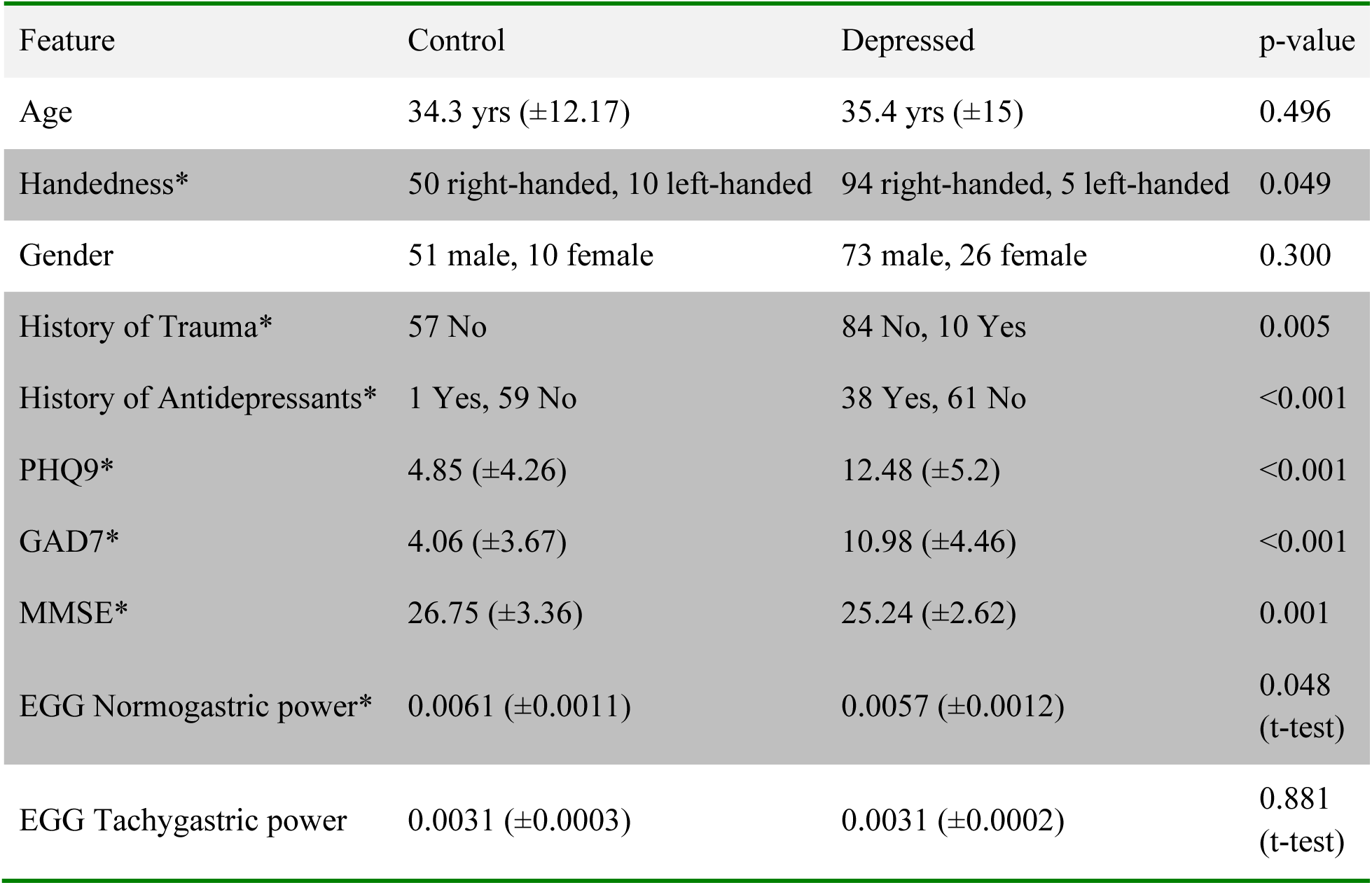
Summary of demographics and baseline electrophysiology in our population from site 1 used for model development. . The statistics indicate the p-value of chi-square tests for Handedness, History of Trauma, History of depression medications, PhQ9, GAD7, MMSE, and t-test of the normogastric EGG power are significantly different between non-clinical (control) and clinical (patient)groups.

The data from site 2 was processed separately using the same methods as those applied to site 1 data. Importantly, the site 2 data served solely as an independent held-out test set to evaluate our model, thereby assessing the robustness and reliability of our model prediction. The age range of the patients from site 2 was 38.7±10 years old, all right handed, 26 males and 19 females, 9 had a history of trauma, 24 had a history of taking antidepressants. Their baseline PhQ9 score was 14.6±5.7, GAD7 was 11.2±4.2, and MMSE was 20.47±5.7.

**Figure 2A** presents the initial patient characteristics from various clinical data collected from visit 1. There were significant differences between control and depressed groups in visit 1 for medicine and trauma history, PHQ9, GAD7, MMSE **(Table 1)**. The clinical participants who showed significant change in mental health (N=28) had a slightly greater mean PHQ9 and GAD7 scores (question-wise) when compared to those who showed no significant change in baseline visit 1 (N=22, **refer to Supplementary figure 1).** Patients who presented significant change in mental health response showed improvement across all questions in PHQ9 and GAD7.

**Figure 2:**
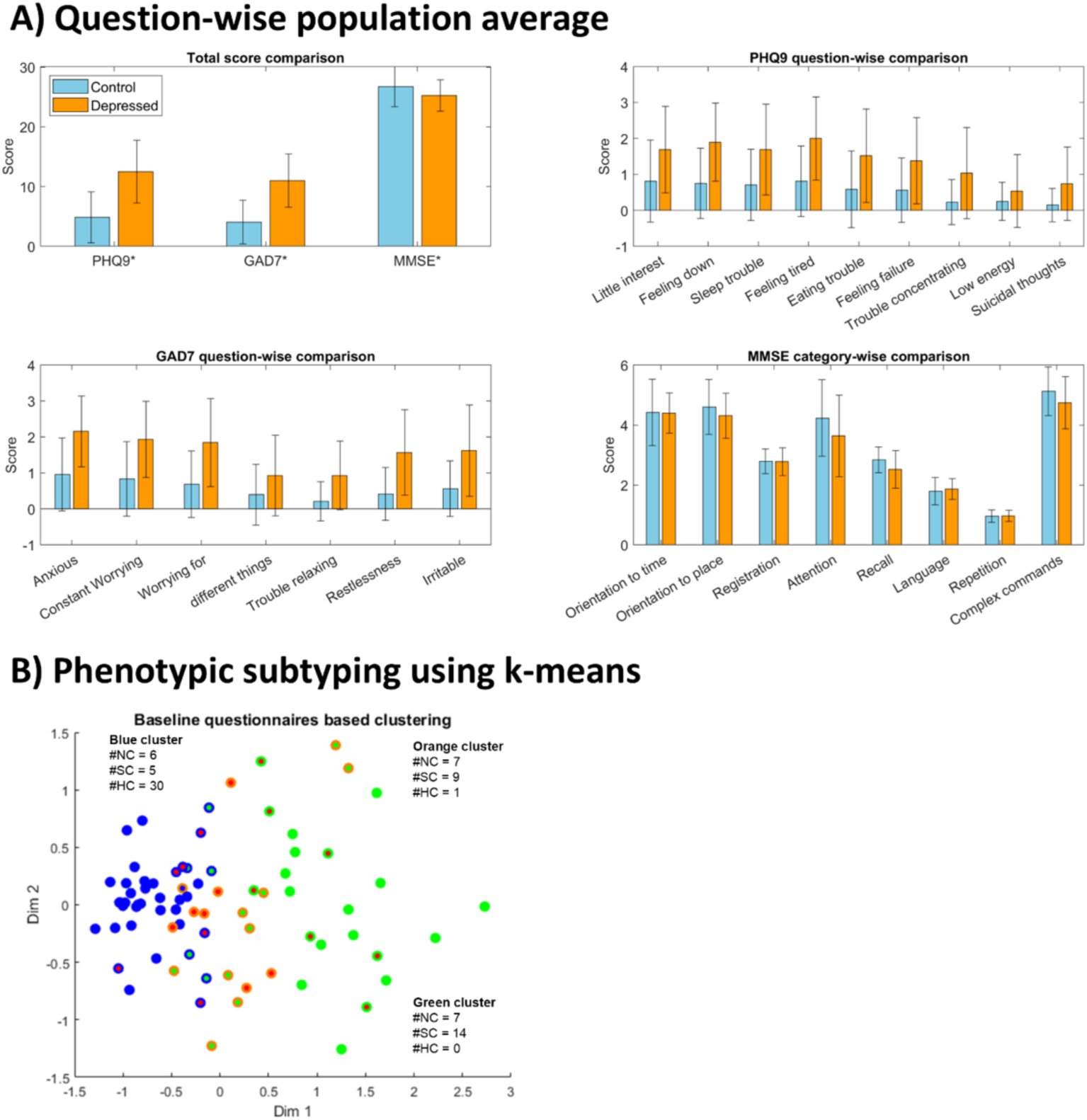
Participants clinical characteristics. **A)** Question-wise comparison of mean scores for the control and depressed populations. **B)** Baseline questionnaire clustering using k-means algorithm: Phenotypic subtyping of information obtained in visit 1 into 3 groups based on their eventual response characteristics in visit 3 (Healthy controls HC, Patients showing significant change in mental health SC, no change in mental health NC), performed using baseline PHQ9, GAD7 and MMSE responses.

**Figure 2B** presents the results of the baseline phenotypic subtyping using just the questionnaires data and we observe that the blue cluster predominantly contained healthy subjects (PHQ9<5, 73%) and the green cluster represent patients showing significant changes in response (67%) and no change in mental health (33%). The orange cluster had almost equal proportion of responders and non-responders. This suggest that based on questionnaires, it is hard to successfully distinguish responders from non-responders and prompts a need to have an objective approach to predict treatment response (N=3 clusters was identified as optimal using elbow and silhouette methods).

### Major theories on frontal activations and brain-gut interactions were sensitive to treatment response

In terms of neurophysiological underpinnings, earlier studies hint at least 4 main theories for predicting treatment response in depression, such as increased theta cordance and increased magnitude of frontal alpha asymmetry in depression. There has been a decrease of excitation inhibition ratio suggesting a plausible role of aperiodic exponent, and an increased gut symptom presentation suggesting a plausible role of gut-brain coupling in patients. We asked whether these four features: gut-brain coupling, aperiodic exponent, theta cordance and frontal alpha asymmetry (right – left) can be reliable markers for our study as well?

At baseline, we did not observe significant differences in theta cordance between control and depressed individuals. However, the feature was predictive of early response fairly in visit 2 during eyes closed cognitive state (d = 0.389, p = 0.007) and were reliable even in visit 3 (d = 0.360, p = 0.043), where the non-responders showed greater reduction in frontal theta cordance when compared to responders.

We also observed increased alpha activations in the right compared to left during baseline in controls than in depressed individuals in resting state eyes open task (d = 0.280, p = 0.003). Interestingly, in eyes closed task (d = 0.344, p = 0.011) the feature was lower in controls, indicating a balancing right to left activations (refer **Figure 3)** and they were early predictive of treatment response especially during interoceptive cognitive state (d = 0.593, p = 0.004).

**Figure 3.**
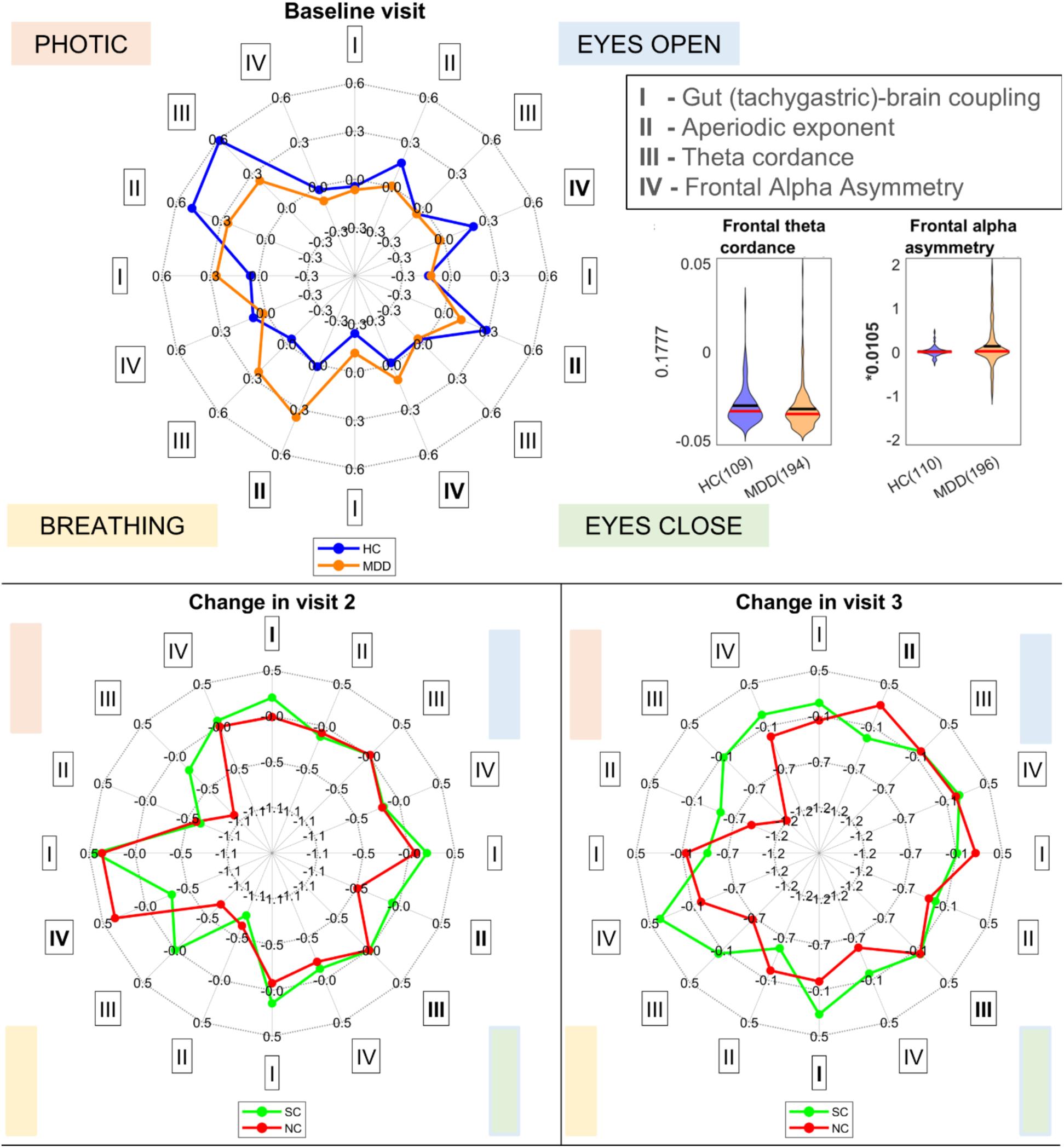
Summary features based on theories of depression. We explore four different theories of depression treatment prediction-increased gut-brain coupling (I), lower aperiodic exponent (II) and lower theta cordance (III), higher frontal alpha asymmetry (IV), when compared to baseline predicts significant response to treatment. Many of these features were sensitive to the cognitive state context to significantly predict the response outcome in patients in baseline visit 1, visit 2 or visit 3. The boldened Greek numerals represent significant differences in the healthy versus patient samples (baseline) or the responders versus non-responders (visits 2, 3) in two sample hypothesis testing, not controlled for multiple comparisons.

The grand average aperiodic exponent across frontal brain regions was significantly more for depressed individuals when compared to healthy participants in breathing task (d = 0.381, p = 0.042). Over time, we noticed the feature decreased more in non-responders especially during eyes closed state (d = 0.328, p = 0.048) in visit 2, but shows the opposite trend in eyes opened state (d = 0.391, p = 0.027) in visit 3.

Although there is no significant difference for gut-brain coupling across tasks at baseline, medication increases the coupling value for responders for eyes open task in visit 2 (d= 0.287, p = 0.029) and breathing task in visit 3 (d = 0.749, p = 0.033).

The above results present evidences against the reliability of any single feature identified in earlier literature, and its predictive ability to classify responders from non-responders. Going forward, we asked whether there are multivariate patterns for response instead of a single biomarker, and whether those response patterns can predict treatment outcome as early by 1 week. To address the questions, we performed machine learning model analysis as detailed in the following section.

### Longitudinal designs and whole-person approach can significantly improve the prediction of responses

How do different forms of data, such as clinical reports, EEG, EGG, collected at different time points of treatment broadly contribute to response prediction? We start investigating this question using a ridge regression model in this section and later in the manuscript through various machine learning models. We setup the simple regression with just the baseline EEG features collected during visit 1. The cross-validated r2 score of the model was 0.4, indicating it explained about 40% variance in response related changes in PhQ9 severity during visit 3 for the patient group. Interestingly, further adding the longitudinal brain information improved the explanation of variance to approximately 60%. And having information about longitudinal changes of brain and gut through time till visit 2 explained >70% of data **(Table 3A)**. Note that if the model has to explain the actual PhQ9 severity in visit 3, baseline alone contributed to about 27%, adding longitudinal EEG explained about 38%, and further adding EGG as well explained 51% variance of absolute severity scores. The above evidence guided us to develop an explainable prediction model that can classify responders from non-responders using longitudinal electrophysiological data.

**Table 3:**
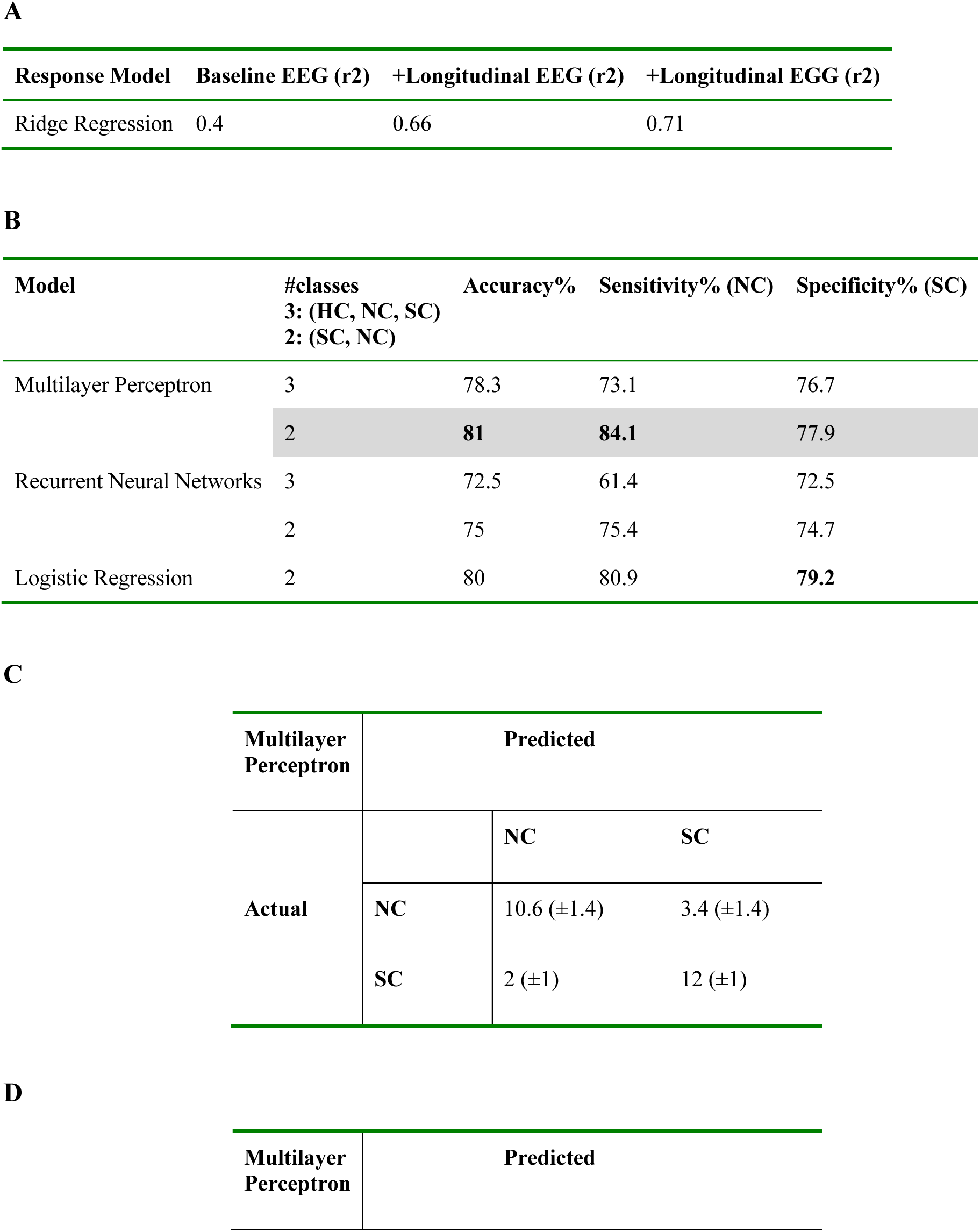
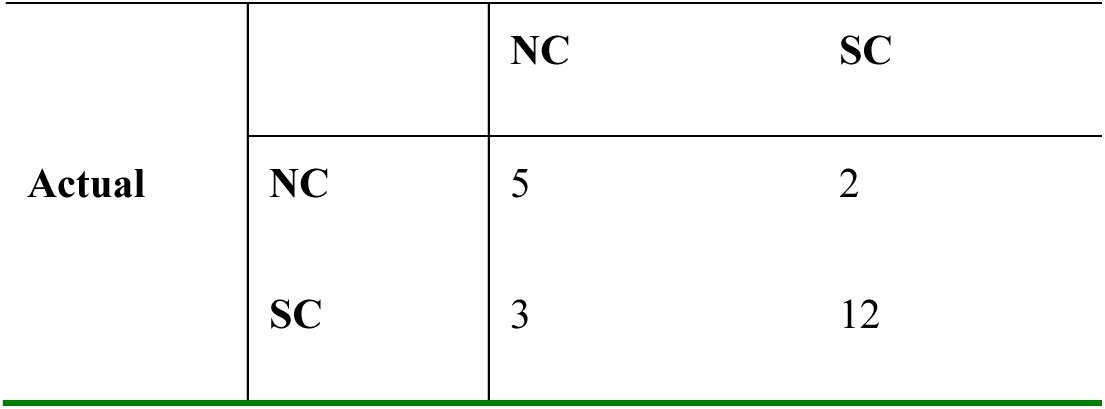
Model selection for predicting treatment outcome. A) Importance of longitudinal design and brain-gut coupling for characterizing treatment strategy of patients: Ridge regression model performance after including various feature groups in steps. **B)** Based on model comparisons, we observe that 2class MLP is best suited for predicting participants with no change in mental health and Logistic Regression is more sensitive for predicting significant change in mental health. C) Confusion matrix from the 2class MLP model analysis predictions on 10 simulations. D) Confusion matrix from the 2class MLP model predictions from independent test data.

| Response Model | Baseline EEG (r2) | +Longitudinal EEG (r2) | +Longitudinal EGG (r2) |
| --- | --- | --- | --- |
| Ridge Regression | 0.4 | 0.66 | 0.71 |

**B**
| Model | #classes<br>3: (HC, NC, SC)<br>2: (SC, NC) | Accuracy% | Sensitivity% (NC) | Specificity% (SC) |
| --- | --- | --- | --- | --- |
| Multilayer Perceptron | 3 | 78.3 | 73.1 | 76.7 |
|  | 2 | **81** | **84.1** | 77.9 |
| Recurrent Neural Networks | 3 | 72.5 | 61.4 | 72.5 |
|  | 2 | 75 | 75.4 | 74.7 |
| Logistic Regression | 2 | 80 | 80.9 | **79.2** |

**C**
| Multilayer Perceptron | Predicted |  |  |
| --- | --- | --- | --- |
|  |  | NC | SC |
| Actual | NC | 10.6 (±1.4) | 3.4 (±1.4) |
|  | SC | 2 (±1) | 12 (±1) |

**D**
| Multilayer Perceptron | Predicted |
| --- | --- |

**Table 3: Model selection for predicting treatment outcome.**
|  |  | NC | SC |
| --- | --- | --- | --- |
| Actual | NC | 5 | 2 |
|  | SC | 3 | 12 |

### A perceptron model can efficiently early predict patients showing significant response to treatment

Next, we asked whether we can early predict the treatment response right at visit 2? Forty eight participants had complete EEG and EGG data for all the tasks of all 3 visits and were included in developing the predictive model **(Figure 1C)**. We used multi-layer perceptron (MLP) and simple recurrent neural network (RNN) models to classify the data for those patients that will exhibit significant reduction of mental health severity (SC), and no change in mental health (NC), controls (HC). The feature selection process resulted in electrophysiological features: 12 (baseline) + 11 (longitudinal) from resting state eyes open task, 10+22 from state eyes closed task, 6+14 in breathing task and the remaining 7+12 while performing photic administration task (refer **Supplementary table 1–** for the feature list), demographics (5) and questionnaires (24). First, we deployed 3-class models, they were validated using k-fold cross validation (k=5) run for 10 instances. The overall cross-validation mean accuracy for MLP was >74% (∼37/48) and for RNN was >70% (34/48) which was well above the chance level of 33% (**Table 3B**).

We also tested two class models specifically trained on patients data, to early predict whether a patient will show significant change/no change in mental health by end of the treatment course. For this question, we compared the performances of the earlier described MLP and RNN architectures but for two class output, along with another Logistic Regression. All three models were trained and cross-validated using 28 subjects from site 1, and performed well above the chance level of 50%. The MLP model outperformed the other 2 with an accuracy of ∼81% (23/28), while the accuracy of the Logistic Regression of ∼79% (22/28) and RNN was ∼75% (21/28). The sensitivity of the 2-class MLP model to predict a participant with no change in mental health was ∼84%. The relative risk of our model, computed as the proportion of subjects falsely predicted as non-responders is 2 subjects (16%). The Matthew’s coefficient accounting for all the depressed subjects is 0.62. The specificity of the model for predicting significant change in mental health is 78% (**Table 3C**). Out of all the 5 individual models and also their nesting architecture, the MLP architecture had a least relative risk, and the model was selected to further explain the importance of each feature towards the prediction using SHAPley values.

Importantly, we also tested using an independent site 2 data, and the MLP model showed accuracy of 77.27%, with sensitivity to non-responders as 71.43% and specificity to response as 80%, confirming the robustness and reliability of our model prediction (**Table 3D)**.

### Feature marginal contribution is sensitive to symptom presentation in patients

We computed Feature importance as the marginal contribution of top feature groups that in sum explained >95% of the observed response prediction (**Figure 4**) in the 2-class multi layer perceptron. The results suggest that overall, changes in left fronto-central absolute theta power, global coherence across bands during photic task and coherence between regions other than central left during eyes closed task are higher for participants showing significant change in mental health. On the other hand, changes in coherence during breathing task, central left coherence during eyes closed task, beta asymmetry between hemispheres (frontal), periodic theta power in frontal left region, aperiodic exponent, and tachygastric gut rhythm-brain coupling decreases.

**Figure 4:**
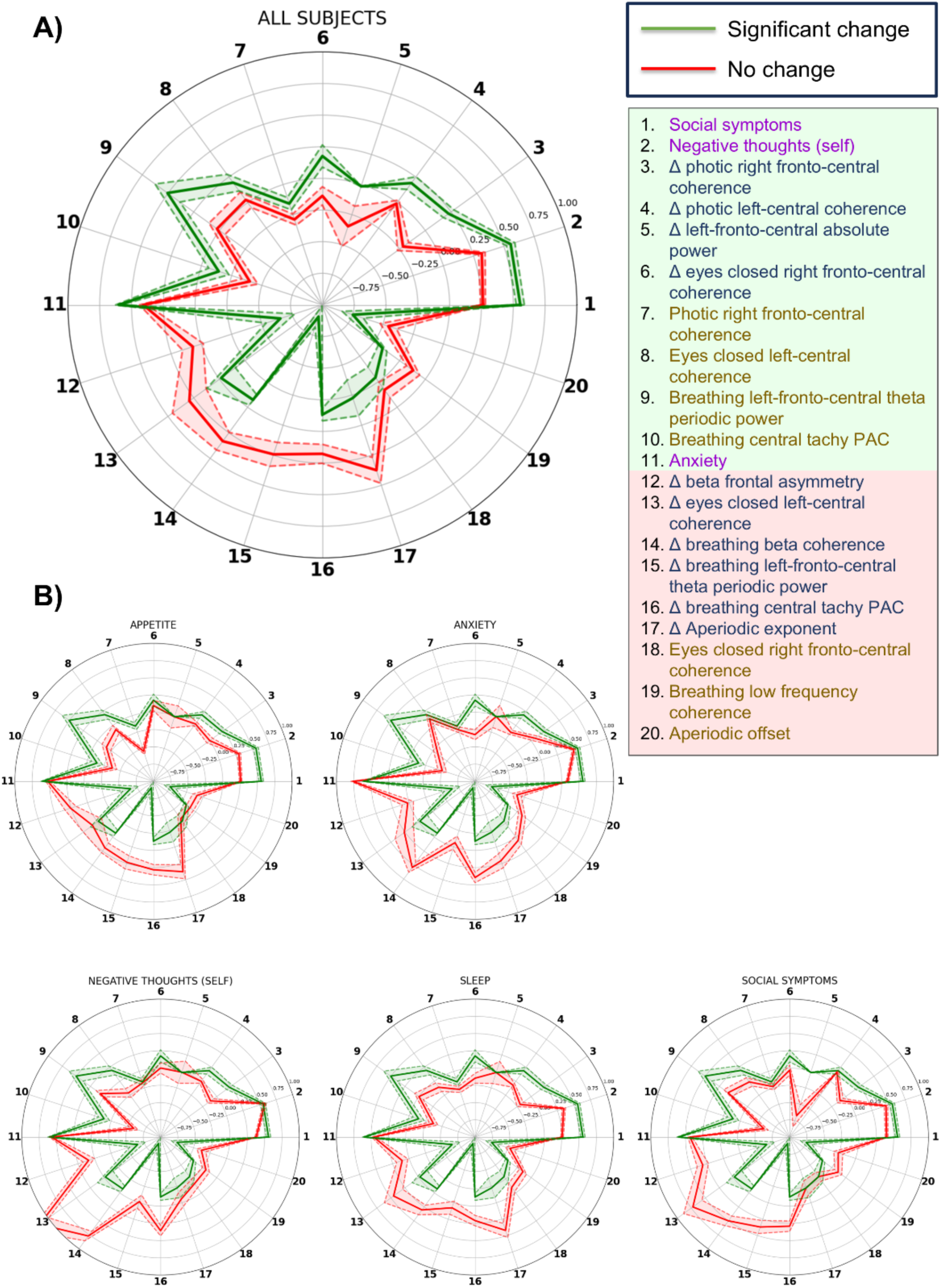
Interpretable and Predictive Modeling of Treatment Outcome. **A)** Zscored normalized comparison between responders (SC-green) and non-responders (NC-red) of top features significantly contributing the multi layer perceptron 2-class model performance. Shaded region represents s.e.m for N=28, the green legend shading reflects feature increase for response while the red indicates the contrary. **B)** The schematic feature profiles for selected participants with high severity in appetite, anxiety, negative thoughts about self, sleep and social symptoms.

**Figure 5:**
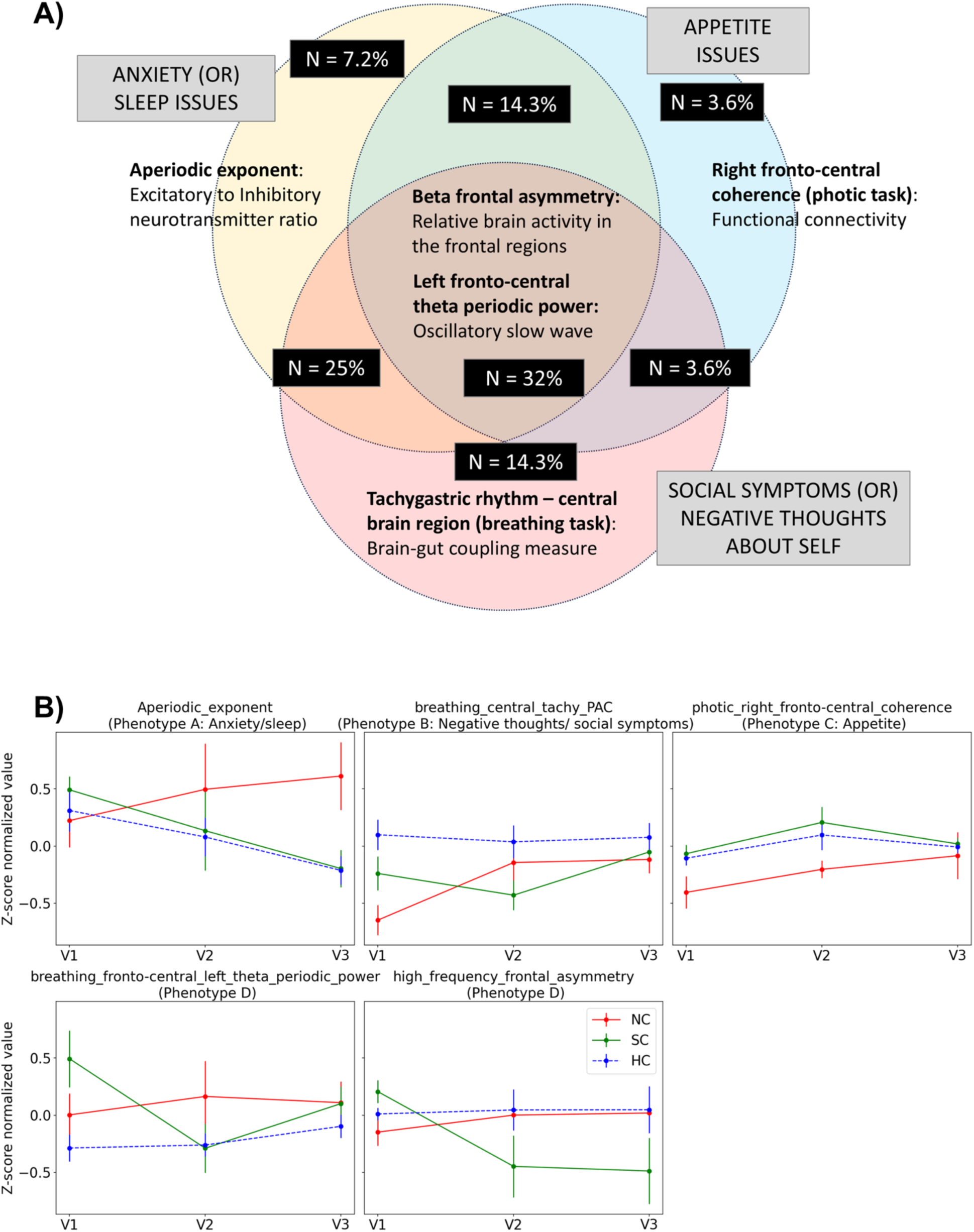
Phenotypic subtype changes through time. **A) Phenotypic subtypes in our patient population represented as a venn diagram.** The diagram shows the proportion of patient population having higher than median severity of the phenotypes indicating dominant sleep or anxiety symptom presentation, gut or homeostatic regulation symptoms, negative thoughts about self-symptoms, and their combinations. (**B) Longitudinal group level changes for top Shapley value features from each phenotype.** The representative features were chosen from Table 4 and the same has been plotted across timepoints (V1, V2, V3) for no change, significant change and healthy controls. The error bars represent s.e.m.

We also observed differences in the features based on the specificity of symptom severity. Longitudinally, in interoceptive breathing task, global beta coherence and tachygastric gut rhythm coupling with central brain regions reduced for responders independent of the symptom manifestation. Aperiodic exponent decreased selectively for responders with high sleep symptom manifestation. Similarly, left fronto-central theta absolute power increased selectively for responders with social symptoms. Longitudinally, decrease in beta frontal asymmetry and left- fronto-central theta periodic power during breathing task were significantly sensitive to treatment response for all symptom groups except anxiety. Overall, we observed different electrophysiological biomarkers to be predictive of treatment outcome depending on their subtype (**Table 4**).

**Table 4:** Significant features for specific phenotypic subtypes-. Electrophysiological feature comparison for responders and non-responders in a subset of population with specific symptom severity, with relatively higher symptom score than the population median. The above table highlights the feature group from the top 20 that are significantly different between the responders and non-responders. The cells highlighted with green has a greater mean for SC than NC and vice versa for red cells, both suggesting that the feature’s response may be linked to the criteria set for a specific phenotypic subtype (column index). The cells depict the mean and std dev of NC and SC datapoints. p-value<0.05 was defined as significant.

|  | Anxiety | Appetite | Negative thoughts (self) | Sleep | Social symptoms |
| --- | --- | --- | --- | --- | --- |
| $\Delta$ photic right fronto-central coherence ($\Delta T_4$ Coh FrCR) | 0.064±0.45,<br>0.181±0.43 | 0.004±0.45,<br>0.284±0.5 | -0.109±0.53,<br>0.131±0.42 | -0.196±0.46,<br>0.222±0.45 | -0.101±0.51,<br>0.174±0.4 |
| $\Delta$ photic left-central coherence ($\Delta T_4$ Coh CL) | -0.123±0.37,<br>0.254±0.52 | 0.029±0.26,<br>0.331±0.5 | -0.001±0.31,<br>0.217±0.55 | -0.07±0.3,<br>0.248±0.52 | 0.161±0.17,<br>0.147±0.51 |
| $\Delta$ left-fronto-central absolute power ($\Delta F_{LC}$ Abs power) | -0.012±0.01,<br>-0.018±0.02 | -0.018±0.01,<br>-0.016±0.02 | -0.012±0.01,<br>-0.016±0.02 | -0.015±0.01,<br>-0.016±0.02 | -0.675±1.93,<br>-0.015±0.02 |
| $\Delta$ eyes closed right-fronto central coherence ($\Delta T_2$ Coh FrCR) | -0.323±0.33,<br>0.377±0.78 | 0.098±0.47,<br>0.237±0.84 | 0.001±0.56,<br>0.293±0.78 | -0.147±0.66,<br>0.273±0.75 | -0.03±0.49,<br>0.216±0.75 |
| Photic right fronto-central coherence ($T_4$ Coh FrCR) | -0.213±0.24,<br>-0.214±0.42 | -0.542±0.29,<br>-0.104±0.37 | -0.171±0.38,<br>-0.216±0.42 | -0.274±0.39,<br>-0.215±0.41 | -0.231±0.36,<br>-0.137±0.42 |
| Eyes closed left-central coherence ($T_2$ Coh CL) | 0.119±0.61,<br>0.259±0.67 | -0.074±0.67,<br>0.345±0.77 | -0.211±0.39,<br>0.263±0.65 | -0.108±0.56,<br>0.241±0.71 | -0.012±0.64,<br>0.264±0.65 |
| Breathing left-fronto-central theta periodic power ($T_3$ FLCL $\theta$ Per power) | -0.34±0.41,<br>0.207±0.89 | -0.16±0.43,<br>0.7±1.15 | 0.072±0.98,<br>0.367±1.1 | 0±0.71,<br>0.398±1.02 | 0.095±0.86,<br>0.521±1.05 |
| Breathing central tachy PAC ($T_3$ Cen-Tachy PAC) | -0.501±0.46,<br>-0.237±0.72 | -0.376±0.62,<br>-0.106±0.82 | -0.593±0.45,<br>-0.186±0.7 | -0.368±0.59,<br>-0.189±0.74 | -0.609±0.45,<br>-0.188±0.66 |
| $\Delta$ beta frontal asymmetry ($\Delta \beta$ Fr Asym) | 0.021±1.17,<br>-0.331±0.81 | 0.259±0.71,<br>-0.612±1.12 | 0.09±1.06,<br>-0.398±0.83 | 0.105±0.83,<br>-0.588±1.04 | 0.315±0.51,<br>-0.48±0.79 |
| $\Delta$ eyes closed left-central coherence ($\Delta T_2$ Coh CL) | 0.261±0.29,<br>0.063±1.24 | 0.087±0.39,<br>0.249±1.33 | 1.154±1.95,<br>0.032±1.26 | 0.453±1.59,<br>0.11±1.2 | 0.723±1.76,<br>-0.106±1.15 |
| **$\Delta$ breathing beta coherence ($\Delta T_3 \beta$ Coh)** | 0.543±0.78,<br>-0.026±0.47 | 0.192±0.12,<br>-0.278±0.33 | 0.772±0.79,<br>-0.054±0.47 | 0.305±0.79,<br>-0.08±0.45 | 0.501±0.56,<br>-0.039±0.41 |
| $\Delta$ breathing left-fronto-central theta periodic power ($\Delta T_3$ FLCL $\theta$ Per power) | -0.066±0.93,<br>-0.586±0.43 | 0.235±0.71,<br>-0.969±0.82 | -0.018±0.84,<br>-0.623±0.47 | 0.078±0.75,<br>-0.825±0.75 | 0.37±0.91,<br>-0.753±0.5 |
| **$\Delta$ breathing central tachy PAC ($\Delta T_3$ Cen-Tachy PAC)** | 0.394±0.74,<br>-0.113±0.4 | 0.282±0.6,<br>-0.176±0.38 | 0.356±0.61,<br>-0.143±0.39 | 0.21±0.67,<br>-0.111±0.42 | 0.291±0.56,<br>-0.13±0.39 |
| $\Delta$ fronto-central Aperiodic exponent ($\Delta Ap$ exponent) | 0.209±1.09,<br>-0.294±1.34 | 0.377±1.03,<br>-0.299±1.41 | -0.049±1.09,<br>-0.214±1.36 | 0.422±1.02,<br>-0.322±1.3 | -0.228±0.68,<br>-0.167±1.24 |
| Eyes closed non-left-central coherence ($T_2$ Coh FrCR) | 0.058±0.38,<br>-0.342±0.76 | -0.33±0.57,<br>-0.054±0.92 | -0.167±0.53,<br>-0.318±0.76 | -0.1±0.66,<br>-0.374±0.72 | -0.29±0.44,<br>-0.251±0.86 |
| Breathing low frequency coherence ($T_3$ Coh $\theta$, $\alpha$) | -0.164±0.5,<br>-0.347±0.37 | -0.348±0.54,<br>-0.416±0.37 | -0.193±0.46,<br>-0.362±0.37 | -0.147±0.54,<br>-0.393±0.37 | -0.135±0.4,<br>-0.383±0.34 |
| Fronto-central Aperiodic offset (Ap offset) | -0.366±0.68,<br>-0.917±0.72 | -0.339±0.56,<br>-0.648±0.79 | -0.381±0.64,<br>-0.94±0.71 | -0.471±0.65,<br>-0.901±0.67 | -0.263±0.45,<br>-0.73±0.75 |

Notably severe anxiety symptoms have shown to have lower remission rate in larger multi- centric International studies (Saveanu et al., 2015). Taking inspiration from RDOC (Research Domain Criteria) for dissociable neural circuit mechanisms as well as the Tripartite theory of Depression (Anna Clark & Watson, 1991), we cluster the phenotype presentations in our participants into gut symptoms or homeostatic regulation issue dominant symptom presentation, cognitive behavioral issues dominant phenotype, and arousal related presentations. They are suggested to have distinct circuit underpinnings to some extent, such as—vagal/gut signals, and cognitive and action execution circuit dynamics, general arousal indicators, respectively.

In particular, we clustered the physiological underpinnings that distinctly represents arousal and anxiety, appetite and homeostatic regulation, self-referential processing and suicidal cognition, and we observed N = 7 being part of pure phenotypic subtypes, and N = 12 presenting a combination of phenotypic subtypes and N = 9 all three-phenotypic subtype presentation. The presence of a symptom was dichotomized based on the median value of the answers to baseline PHQ9 questions (refer to Supplementary material section 3 for details of the exact questions that were averaged to compute the subject’s symptom score). Participants with scores greater than the median value were considered to manifest the symptom.

For participants with anxiety or sleep issue, the aperiodic exponent significantly decreases across visits for responders (SC) while it remains the same for non-responders (NC) (p-value < 0.001). When similar analysis was performed for participants with negative thoughts about their self, we observed that the representative feature, i.e., coupling between tachygastric gut rhythm and central brain regions, shows sudden increase for non-responders post medication (p-value = 0.006) and this remains consistently high across timepoints. For participants exhibiting appetite or homeostatic dysregulation symptoms, the coherence in right fronto-central regions during photic administration task remains consistently high from baseline (p-value = 0.042) to visit 2 (p-value = 0.031) for responders when compared to non-responders. Notably, the significant responder profile closely follows the expected change in control population. Participants exhibiting composite symptom presentation show decrease in left fronto-central theta periodic power (p -value = 0.003) and beta frontal asymmetry (p-value = 0.015) for responders. The reduction in beta frontal asymmetry is consistently seen across timepoints for responders, however was not observed in the healthy controls (p-value = 0.013).

## DISCUSSION

### Current “Watchful Waiting” Treatment Approaches

Treatments for depression, such as medication, supportive therapies, and interventions like magnetic stimulation or cognitive restructuring, rely heavily on continuous monitoring to predict treatment outcomes. However, with the increasing patient-to-clinician ratio, accurately predicting treatment efficacy through traditional in-person interviews has become increasingly difficult. Over a third of patients discontinue treatment within the first 30 days due to these intolerable side effects. If depression persists after trying two different medications, the patient is often deemed treatment-resistant. The “watchful waiting” approach, in which clinicians wait and observe for weeks to determine if a treatment is effective, has been criticized for its inefficiency and high resource consumption. Studies show that less than 20% of patients benefit from this method (Bauer et al., 2007; Bockting et al., 2008; Fochtmann & Gelenberg, 2005; Kas et al., 2025; Pigott et al., 2010; Quitkin et al., 2003). Given these challenges, there is an urgent need for tools that can predict treatment outcomes sooner—within 2 weeks—enabling clinicians to adjust strategies before patients experience prolonged suffering or side effects.

Our study addresses this gap by showing that early predictions of treatment outcomes in depression can be made within just two weeks by analyzing electrophysiological markers in both the brain and gut, with 77.27% test accuracy. Specifically, our study identifies early markers that were significantly predictive of treatment response, as early as in 7-10 days from the start of the treatment. Gut-brain connectivity, a novel aspect of our approach, provided additional early markers, pointing to the utility of incorporating gut dysfunctions alongside brain measures. These findings underscore the importance of integrating whole-body physiology in treatment predictions, which allows clinicians to intervene much earlier in the treatment process.

An important aspect of our study is the use of phenotypic subtyping—identifying depression subtypes based on symptom profiles (e.g., anxiety, sleep disturbances, appetite issues, negative self-thoughts, or social withdrawal)—to predict treatment outcomes. Our results suggest that personalized predictive markers should be tailored to each subtype for more accurate and efficient outcomes. For instance, we found that increased fronto-central excitation-inhibition ratios were the strongest predictors of treatment response for patients with dominant anxiety or sleep-related symptoms. In contrast, decreased tachygastric coupling predicted a positive response for patients with negative self-thoughts. Additionally, right fronto-central connectivity was associated with positive treatment outcomes in patients experiencing appetite-related issues.

### Both periodic and aperiodic activations predict treatment outcome

Frontal changes in various bands have been markers of depressive symptoms and its treatment response according to numerous studies. Some specific markers include increased theta cordance in depressed individuals especially in the frontal region (de la Salle et al., 2020). However, there has been contradictory viewpoints on the oscillatory mechanistic underpinnings because when separating the theta power into its aperiodic and periodic components, the aperiodic one reflected the depression measures relatively (Smith et al., 2023). These contradicting evidences question the very processes mediating the severity of depression and their responses to intervening treatment. We teased apart the periodic components of oscillation to specifically understand the role of periodic theta components in predicting response to medicine. Interestingly, periodic theta power in the left frontal and central region reduced post medication as mentioned in previous literature. On the other hand, aperiodic parameters such as slope and offset played a crucial role in categorizing someone as a responder/non-responder highlighting the importance of using aperiodic parameters as a biomarker for treatment outcome prediction. Aperiodic offset and aperiodic exponent are a measure of global signal power and excitation-inhibition E/I neurotransmitter ratio. A greater offset and a lower exponent value indicates balanced power between bands and increased global power. A steeper exponent indicates greater lower frequency power when compared to higher frequency power and is commonly observed in state of relaxation. Antidepressants such as SSRI, SNRI are said to increase the global band power of the signal.

### Frontal beta asymmetry can early predict treatment outcome

The slowness of brain activity, in other words the power of lower frequency spectrum was related to treatment response by many studies even in alpha band. The frontal alpha asymmetry between the left and right hemispheres were in particular a signature of depression severity. Studies suggest that asymmetry in the brain facilitated a differentiating response to positive versus negative affect, with more attenuation to positive affect and increased response to negative affect (Davidson, 2003; Harmon-Jones et al., 2010). However, the frontal alpha asymmetry was not consistently observed to be indicative of treatment response, and rather as a marker of depression experience (van der Vinne et al., 2019). The positive or negative affective stimulus induced emotional responses were also separately studied by few scientists, to investigate the elicited emotional sensitivity to depression severity and treatment response, but there weren’t any robust results (Kaviani et al., 2004; Kołodziej et al., 2021). In our study, we did not observe strong effect of the frontal alpha power for predicting response. Interestingly, we observed frontal beta asymmetry to be sensitive to treatment response and the beta power in the frontal left region relative to the right region increased significantly for depressed individuals who responded to medication. Some studies specifically looked into reward processing and inhibitory control mechanisms for understanding the severity (Oh et al., 2023; Yitzhak et al., 2023) for response to treatment (Brandt et al., 2021): Not-surprisingly, evidences point to the significant presence of reward information in the frontal beta oscillations, suggesting their potential role for early predicting of outcome (Koloski et al., 2024).

### Brain connections and its gut modulatory indices hold important information on treatment effects

Recently, the role of connectivity has garnered more attention (Elam et al., 2021), and a reduced connectivity were found in people experiencing depression (Huang et al., 2023; Roemer et al., 1992). More structural level investigation also suggested the possibility of a short-range excitation and long-range reduction in connections in depression (Fingelkurts et al., 2007). In our study, we find a significant contribution of the connectivity features, in all the theta, alpha and beta bands, for predicting the treatment response.

Our study for the first time tests the role of gut-brain coupling in explaining depression severity and response to treatment. Interestingly, we find it to be predicting the response to treatment as early as within two weeks from the treatment onset. The role of gut dysfunction in depression has been investigated by numerous studies, however to our understanding, the gut dysfunction exhibited as abnormal motility of the stomach and the intestine has not been explored to strategizing the treatment for depression. Our study strongly suggests that looking into the gut-brain coupling can assist with personalizing treatment strategy in an optimal fashion. Importantly, it highlights that a scalable electrogastrography tool could be used for the purpose of computing the gut-brain coupling. In our population, we observed that the brain-gut coupling in tachygastric frequency reduces significantly for responders because of medication.

Furthermore, our study also proposes that longitudinal design that can track the plasticity in neural circuits are able to explain the depression measures better than the cross-sectional design of studying and developing of prediction models based on the baseline time sample in silo. This is in lines with many studies that find greater sensitivity for a longitudinal marker in depression (Bares et al., 2015; Schwartzmann et al., 2024).

### Cognitive Control and Sensitivity of Task Paradigms in Treatment Response

An important finding in our study is the sensitivity of cognitive control during the administration of specific task paradigms, which serve as reliable markers of treatment response. Beyond traditional resting-state paradigms (eyes open and eyes closed), we identified highly predictive markers during photic flicker presentation and hyperventilating breathing tasks. The photic flicker effect, which is known to modulate cognitive processes depending on the frequency of the stimulus, can influence how external stimuli or internal thoughts are processed, especially through phase coupling with the flicker onset (Thut et al., 2011; Williams et al., 2006). Similarly, deep breathing paradigms, which have previously been shown to be sensitive to depression symptoms, were also found to be valuable for assessing treatment response in our study (Segal et al., 2002; Michalak et al., 2010).

### Phenotype-Based Analysis of Treatment Response Mechanisms

Analyzing phenotype-based features for treatment response offers valuable insights into the distinct neurophysiological mechanisms underlying each depression subtype. Our findings identify specific markers for each subtype, which can inform more targeted treatment strategies.

### Phenotype A - Anxiety/Sleep

Dysfunction of the Locus coeruleus (LC) has been implicated in depression, leading to reduced norepinephrine in the system, which plays a key role in the sleep/wake cycle and arousal (Gong et al., 2021; Grimm et al., 2024). Electrophysiologically, LC dysfunction manifests as reduced coherence, increased aperiodic exponent, and altered Excitatory/Inhibitory (E/I) neurotransmitter ratios. In participants with dominant anxiety and sleep issues, we observe reduced coherence during the photic and eyes-closed tasks, which increases after treatment. Additionally, the aperiodic exponent in the fronto-central region decreases, indicating a shift toward a more balanced E/I ratio. We propose that interventions (ex. Lima et al., 2023) regulating the overall fronto-central E/I ratio may specifically help in addressing this subtype related symptoms.

### Phenotype B - Negative thoughts about self / social symptoms

Depression significantly impairs self-referential cognition, which is closely linked to the default mode network (DMN), a network active during resting states and involved in processing self-related information (Chou et al., 2023). Hyperactivity of the DMN is often associated with rumination, a hallmark of depression where individuals engage in negative self-reflection (Liston et al., 2014). In our study, participants with negative self-thoughts or social symptoms showed increased left fronto-central power after treatment, which we interpret as a potential marker of DMN regulation. Moreover, brain-gut coupling, particularly in the somatosensory-motor region during interoceptive breathing tasks, was significantly disrupted in these individuals but improved post-treatment. Interestingly earlier studies support somato-sensory motor sensitivity and their activations to be a significant marker of intervention for suicidal thoughts (Kerr et al., 2013). We propose that intervention should specifically focus to decrease the activity of DMN (ex. rTMS, Liston et al., 2014), and improve the sensory-motor activations to interoceptive signals, to address this subtype with increased negative thoughts about oneself.

### Phenotype C – Appetite

The insula plays a central role in regulating gastric motility and receives afferent inputs from the nucleus tractus solitarius (NTS), which is involved in processing visceral sensory information from the vagus nerve (Travagli & Anselmi, 2016). Dysfunction in these regions can result in irregular gastric activity, often manifesting as appetite issues in depression. In our study, we found decreased right fronto-central connectivity in participants with appetite-related symptoms during photic stimulation, which may be linked to underlying insular dysfunction. The NTS, which is responsible for glutamate release and relaying vagus nerve signals, may be central to this issue. We propose that vagus nerve stimulation could be an effective treatment for individuals with this phenotype, improving homeostatic regulation and appetite-related symptoms.

### Phenotype D - Composite symptom profile

For patients with a composite symptom profile, where symptoms span appetite, anxiety, sleep disturbances, and negative thoughts, we observed highly sensitive markers during interoceptive breathing tasks. Theta periodic power in the left fronto-central region was elevated at baseline but decreased after treatment in responders. Additionally, frontal beta asymmetry, with increased activity in the left frontal region, served as a key predictor of treatment response. Given the complex nature of this phenotype, a multimodal treatment strategy—drawing from the approaches used for the more pure subtypes—may be most effective for addressing the combined symptoms presented in this group.

### Check for Information leakage in our methods

The features that significantly explained the response across the entire dataset were highly correlated with those observed in a subset comprising 80% of the population (Pearson *corr* = 0.9), ruling out information leakage in our results. Additionally, we ensure that the training and test datasets do not overlap and employ k-fold cross-validation approach to further validate our findings.

### In summary

Overall, our study demonstrates that integrating brain and gut data—along with phenotypic subtyping—can reliably predict a patient’s likelihood of responding to treatment within just two weeks. This is a significant improvement over the standard 4-6 week assessment period, offering clinicians the ability to adapt treatment strategies more quickly and effectively. By understanding the relationship between neural and gut circuits in different phenotypic subtypes, we can provide a personalized, precision medicine approach that aligns treatment strategies with specific symptom profiles, ultimately improving patient outcomes and minimizing unnecessary side effects.

Our study has many limitations. Our sample was limited to a part of India. The study does not account for the role of genetics (Shadrina et al., 2018), social and cultural factors (Kupferberg & Hasler, 2023) influencing the onset and progression of mental health issues, and designing of treatment strategies. As the study was controlled in a real-world clinical setting, as experimenters, we had no control over the treatment strategy. Our future plan is to extend the study to investigating multiple national and international sites for validation of the identified markers of prediction. Moreover, many clinicians administer multimodal treatment, that is medicine in combination with alternative treatments, repetitive transcranial magnetic stimulation, as an intervention. Our future work will also include understanding of the physiological differences due to treatment modality for predicting outcome. Further, we aim to extend the study with high resolution data acquisition of the brain and gut signals to understand the response cortical source dynamics as well as the gut wave propagation in the stomach and intestine.

## Data Availability

All data produced in the present study are available upon reasonable request to the authors

## Acknowledgements

We thank Jyoti Mishra, Dhakshin Ramanathan, Venkatasubramanian, Srinivasa Chakravarthy, for some insightful discussions.

## Data Availability

The datasets generated and/or analyzed during the current study are not publicly available due to the sensitive nature of the clinical and electrophysiological data and to protect participant privacy, in accordance with institutional ethics approval. However, appropriate data will be provided upon request to the corresponding author.

## REFERENCE

1. Arevalo-Rodriguez, I., Smailagic, N., i Figuls, M. R., Ciapponi, A., Sanchez-Perez, E., Giannakou, A., Pedraza, O. L., Cosp, X. B., & Cullum, S. (2015). Mini-Mental State Examination (MMSE) for the detection of Alzheimer’s disease and other dementias in people with mild cognitive impairment (MCI). Cochrane Database of Systematic Reviews, 3.

2. Athreya, A. P., Brückl, T., Binder, E. B., John Rush, A., Biernacka, J., Frye, M. A., Neavin, D., Skime, M., Monrad, D., Iyer, R. K., Mayes, T., Trivedi, M., Carter, R. E., Wang, L., Weinshilboum, R. M., Croarkin, P. E., & Bobo, W. V. (2021). Prediction of short-term antidepressant response using probabilistic graphical models with replication across multiple drugs and treatment settings. Neuropsychopharmacology, 46(7), 1272–1282. 10.1038/s41386-020-00943-x

3. Bares, M., Novak, T., Kopecek, M., Brunovsky, M., Stopkova, P., & Höschl, C. (2015). The effectiveness of prefrontal theta cordance and early reduction of depressive symptoms in the prediction of antidepressant treatment outcome in patients with resistant depression: Analysis of naturalistic data. European Archives of Psychiatry and Clinical Neuroscience, 265(1), 73–82. 10.1007/s00406-014-0506-8

4. Bauer, M., Bschor, T., Pfennig, A., Whybrow, P. C., Angst, J., Versiani, M., & Möller, H. J. (2007). World federation of societies of biological psychiatry (WFSBP) guidelines for biological treatment of unipolar depressive disorders in primary care. World Journal of Biological Psychiatry, 8(2), 67–104. 10.1080/15622970701227829

5. Bockting, C. L. H., ten Doesschate, M. C., Spijker, J., Spinhoven, P., Koeter, M. W. J., Schene, A. H., & DELTA study group. (2008). Continuation and maintenance use of antidepressants in recurrent depression. Psychotherapy and Psychosomatics, 77(1), 17–26. 10.1159/000110056

6. Cao, Z., Lin, C.-T., Ding, W., Chen, M.-H., Li, C.-T., & Su, T.-P. (2019). Identifying Ketamine Responses in Treatment-Resistant Depression Using a Wearable Forehead EEG. IEEE Transactions on Biomedical Engineering, 66(6), 1668–1679. IEEE Transactions on Biomedical Engineering. 10.1109/TBME.2018.2877651

7. Chou, et al., "The default mode network and rumination in individuals at risk for depression", Social Cognitive and Affective Neuroscience, January 2023, 18(1), 10.1093/scan/nsad032

8. Clark and Watson, "516-336 & Heninger", Journal of Abnormal Psychology, 1991, 100(3), Publisher: Maser & Cloninger

9. Donoghue, T., Haller, M., Peterson, E. J., Varma, P., Sebastian, P., Gao, R., Noto, T., Lara, A. H., Wallis, J. D., & Knight, R. T. (2020). Parameterizing neural power spectra into periodic and aperiodic components. Nature Neuroscience, 23(12), 1655–1665.

10. Drossman, D. A. (2016). Functional gastrointestinal disorders: History, pathophysiology, clinical features, and Rome IV. Gastroenterology, 150(6), 1262–1279.

11. Fochtmann, L. J., & Gelenberg, A. J. (2005). Guideline Watch: Practice Guideline for the Treatment of Patients With Major Depressive Disorder, 2nd Edition. FOCUS, *3*(1), 34–42. 10.1176/foc.3.1.34

12. Gautam, et al. Clinical Practive Guidelines for the management of Depression, Indian Journal of Psychiatry, 2017, 10.4103/0019-5545-196973

13. Gong, et al., "The Abnormal Functional Connectivity in the Locus Coeruleus-Norepinephrine System Associated With Anxiety Symptom in Chronic Insomnia Disorder", Frontiers in Neuroscience, May 2021, 15, Article 678465, 10.3389/fnins.2021.678465

14. Goyal, O., Nohria, S., Dhaliwal, A. S., Goyal, P., Soni, R. K., Chhina, R. S., & Sood, A. (2021). Prevalence, overlap, and risk factors for Rome IV functional gastrointestinal disorders among college students in northern India. Indian Journal of Gastroenterology, 40(2), 144–153. 10.1007/s12664-020-01106-y

15. Grimm, et al., "Tonic and burst-like locus coeruleus stimulation distinctly shift network activity across the cortical hierarchy", Nature Neuroscience, November 2024, 27, 2167– 2177, 10.1038/s41593-024-01755-8

16. Hamilton, M. (1960). The Hamilton Depression Scale—Accelerator or break on antidepressant drug discovery. Psychiatry, 23, 56–62.

17. Hasanzadeh, F., Mohebbi, M., & Rostami, R. (2019). Prediction of rTMS treatment response in major depressive disorder using machine learning techniques and nonlinear features of EEG signal. Journal of Affective Disorders, 256, 132–142. 10.1016/j.jad.2019.05.070

18. Jaworska, N., de la Salle, S., Ibrahim, M.-H., Blier, P., & Knott, V. (2019). Leveraging Machine Learning Approaches for Predicting Antidepressant Treatment Response Using Electroencephalography (EEG) and Clinical Data. Frontiers in Psychiatry, 9. 10.3389/fpsyt.2018.00768

19. John Rush, A., Trivedi, M. H., Wisniewski, S. R., Nierenberg, A. A., Stewart, J. W., Warden, D., George Niederehe, M., Thase, M. E., Lavori, P. W., Lebowitz, B. D., McGrath, P. J., Rosenbaum, J. F., Sackeim, H. A., Kupfer, D. J., Luther, J., & Maurizio Fava, M. (2006). Acute and Longer-Term Outcomes in Depressed Outpatients Requiring One or Several Treatment Steps: A STAR*D Report (Am J Psychiatry, Vol. 163, Issue 11, pp. 1905–1917). www.star-d.org

20. Keitner, G. I., Solomon, D. A., & Ryan, C. E. (2008). STAR*D: Have We Learned the Right Lessons? American Journal of Psychiatry, 165(1), 133–133. 10.1176/appi.ajp.2007.07081375

21. Kerr, C. E., Sacchet, M. D., Lazar, S. W., Moore, C. I., & Jones, S. R. (2013). Mindfulness starts with the body: Somatosensory attention and top-down modulation of cortical alpha rhythms in mindfulness meditation. Frontiers in Human Neuroscience, 7, Article 12. 10.3389/fnhum.2013.00012

22. Knott, V., Mahoney, C., Kennedy, S., & Evans, K. (2001). EEG power, frequency, asymmetry and coherence in male depression. Psychiatry Research: Neuroimaging, 106(2), 123–140.

23. Kołodziej, A., Magnuski, M., Ruban, A., & Brzezicka, A. (2021). No relationship between frontal alpha asymmetry and depressive disorders in a multiverse analysis of five studies. eLife, 10, e60595. 10.7554/eLife.60595

24. Kroenke, K., Spitzer, R. L., & Williams, J. B. (2001). The PHQ-9: Validity of a brief depression severity measure. Journal of General Internal Medicine, 16(9), 606–613.

25. Kubo, et al., "Predicting relapse from the time to remission during the acute treatment of depression: A re-analysis of the STAR*D data", Journal of Affective Disorders, January 2023, 320, 710–715, 10.1016/j.jad.2022.09.162

26. Lee, H. S., Baik, S. Y., Kim, Y.-W., Kim, J.-Y., & Lee, S.-H. (2020). Prediction of Antidepressant Treatment Outcome Using Event-Related Potential in Patients with Major Depressive Disorder. Diagnostics, 10(5), Article 5. 10.3390/diagnostics10050276

27. Leuchter, A. F., Cook, I. A., Hunter, A. M., & Korb, A. S. (2009). *The current treatment paradigm for Major Depressive Disorder A new paradigm for the prediction of antidepressant treatment response* (Dialogues Clin Neurosci, Vol. 11, pp. 435–446). www.dialogues-cns.org

28. Lieber, A. L., & Prichep, L. S. (1988). Diagnosis and subtyping of depressive disorders by quantitative electroencephalography: I. Discriminant analysis of selected variables in untreated depressives. The Hillside Journal of Clinical Psychiatry, 10(1), 71–83.

29. Lima, et al., "Acute physical exercise improves recognition memory via locus coeruleus activation but not via ventral tegmental area activation", Physiology and Behavior, December 2023, 272, 114370, 10.1016/j.physbeh.2023.114370

30. Liston, et al., "Default mode network mechanisms of transcranial magnetic stimulation in depression", Biological Psychiatry, 2014, 76(7), 517–526, 10.1016/j.biopsych.2014.01.023

31. Lydiard, R. B. (2001). Irritable bowel syndrome, anxiety, and depression: What are the links? Journal of Clinical Psychiatry, 62, 38–47.

32. Masand, P. S., Kaplan, D. S., Gupta, S., Bhandary, A. N., Nasra, G. S., Kline, M. D., & Margo, K. L. (1995). Major depression and irritable bowel syndrome: Is there a relationship? The Journal of Clinical Psychiatry, 56(8), 363–367.

33. Michalak, J., Troje, N. F., & Heidenreich, T. (2010). Embodied effects of mindfulness- based cognitive therapy. Journal of Psychosomatic Research, 68, 311–314.

34. Overs, J., Morgan, S., Apputhurai, P., Tuck, C., & Knowles, S. R. (2024). Comparing the prevalence and association between anxiety, depression and gastrointestinal symptoms in gastroparesis versus functional dyspepsia: A systematic review and meta-analysis. Journal of Psychosomatic Research, 183, 111834. 10.1016/j.jpsychores.2024.111834

35. Pigott, H. E., Leventhal, A. M., Alter, G. S., & Boren, J. J. (2010). Efficacy and effectiveness of antidepressants: Current status of research. Psychotherapy and Psychosomatics, 79(5), 267–279. 10.1159/000318293

36. Quitkin, F. M., Petkova, E., McGrath, P. J., Taylor, B., Charles Beasley, Mp., Stewart, J., Amsterdam, J., Fava, M., Rosenbaum, J., Reimherr, F., Fawcett, J., Chen, Y., & Klein, D. (2003). Article When Should a Trial of Fluoxetine for Major Depression Be Declared Failed? (Am J Psychiatry, Vol. 160, pp. 4–4). http://ajp.psychiatryonline.org

37. Sadat Shahabi, M., Shalbaf, A., & Maghsoudi, A. (2021). Prediction of drug response in major depressive disorder using ensemble of transfer learning with convolutional neural network based on EEG. Biocybernetics and Biomedical Engineering, 41(3), 946–959. 10.1016/j.bbe.2021.06.006

38. Saveanu, et al., "The International Study to Predict Optimized Treatment in Depression (iSPOT-D): Outcomes from the acute phase of antidepressant treatment", Journal of Psychiatric Research, February 2015, 61, 1–12, 10.1016/j.jpsychires.2014.12.018

39. Segal, Z. V., Williams, J. M., & Teasdale, J. D. (2002). Mindfulness-based cognitive therapy for depression–A new approach to preventing relapse. New York: Guilford Press.

40. Smith, S. E., Ma, V., Gonzalez, C., Chapman, A., Printz, D., Voytek, B., & Soltani, M. (2023). Clinical EEG slowing induced by electroconvulsive therapy is better described by increased frontal aperiodic activity. Translational Psychiatry, 13(1), 1–10. 10.1038/s41398-023-02634-9

41. Spitzer, R. L., Kroenke, K., Williams, J. B., & Löwe, B. (2006). A brief measure for assessing generalized anxiety disorder: The GAD-7. Archives of Internal Medicine, 166(10), 1092–1097.

42. Tennant, R., Hiller, L., Fishwick, R., Platt, S., Joseph, S., Weich, S., Parkinson, J., Secker, J., & Stewart-brown, S. (2007). The Warwick-Edinburgh Mental Well-being Scale (WEMWBS): Development and UK validation. Health and Quality of Life Outcomes, 5, 1–13. 10.1186/1477-7525-5-63

43. Travagli and Anselmi, "Vagal neurocircuitry and its influence on gastric motility", Nature Reviews Gastroenterology and Hepatology, July 2016, 13, 389–401, 10.1038/nrgastro.2016.76

44. Trivedi, M. H., John Rush, A., Wisniewski, S. R., Nierenberg, A. A., Warden, D., Louise Ritz, M., Grayson Norquist, M., Howland, R. H., Lebowitz, B., McGrath, P. J., Shores-Wilson, K., Biggs, M. M., Balasubramani, G., & Fava, M. (2006). Article Evaluation of Outcomes With Citalopram for Depression Using Measurement-Based Care in STAR*D: Implications for Clinical Practice STAR*D Study Team (Am J Psychiatry, Vol. 163, pp. 1– 1). http://ajp.psychiatryonline.org

45. van der Vinne, N., Vollebregt, M. A., van Putten, M. J. A. M., & Arns, M. (2019). Stability of frontal alpha asymmetry in depressed patients during antidepressant treatment. NeuroImage: Clinical, 24, 102056. 10.1016/j.nicl.2019.102056

46. Warden, D., Madhukar Trivedi, M. H., Wisniewski, S. R., Davis, L., Nierenberg, A. A., Gaynes, B. N., Sidney Zisook, M., Hollon, S. D., Balasubramani, G., Howland, R., Fava, M., Stewart, J. W., & John Rush, A. (2007). Predictors of Attrition During Initial (Citalopram) Treatment for Depression: A STAR*D Report (Am J Psychiatry, Vol. 164).

47. Whitton, et al., "Pretreatment Rostral Anterior Cingulate Cortex Connectivity With Salience Network Predicts Depression Recovery: Findings From the EMBARC Randomized Clinical Trial", Biological Psychiatry, May 2019, 85(10), 872–880, 10.1016/j.biopsych.2018.12.007

48. Zamani, M., Alizadeh-Tabari, S., & Zamani, V. (2019). Systematic review with meta-analysis: The prevalence of anxiety and depression in patients with irritable bowel syndrome. Alimentary Pharmacology & Therapeutics, 50(2), 132–143.

