## Supplementary material for "Subtyping Depression using Brain-Gut Electrophysiology for Early Prediction of Antidepressant Response: a multicentric clinical study"

SUPPLEMENTARY MATERIALS

1) ***Questionnaires question-wise differences for responders vs non-responders***

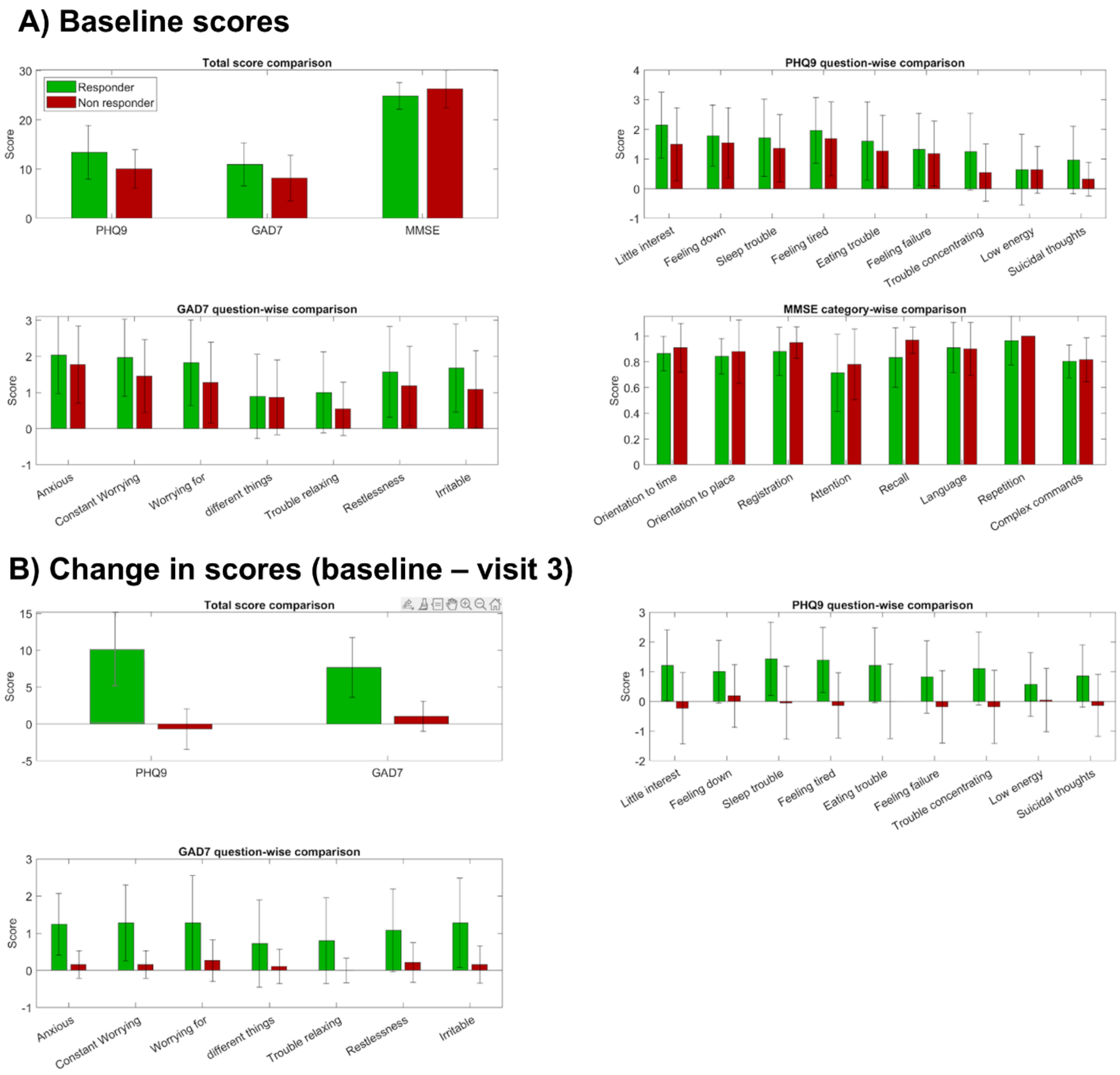


Figure I: Question-wise comparison for responders and non-responders.

From the above figure, we observe that there is no significant difference between the scores of responders and non-responders at the baseline. Post treatment, responders show significant decrease in severity across all symptoms when compared to non-responders.

*Elbow and silhouette scores for baseline questionnaire-based k-means clustering*

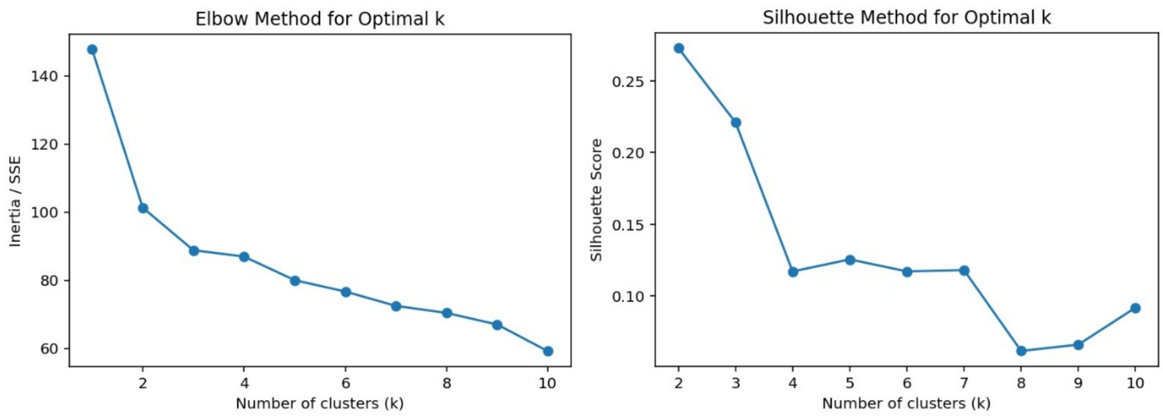


As we needed at least 3 clusters, we set the value of k to be 3 based on the above results.

### 2) *Feature comparison between participant groups*

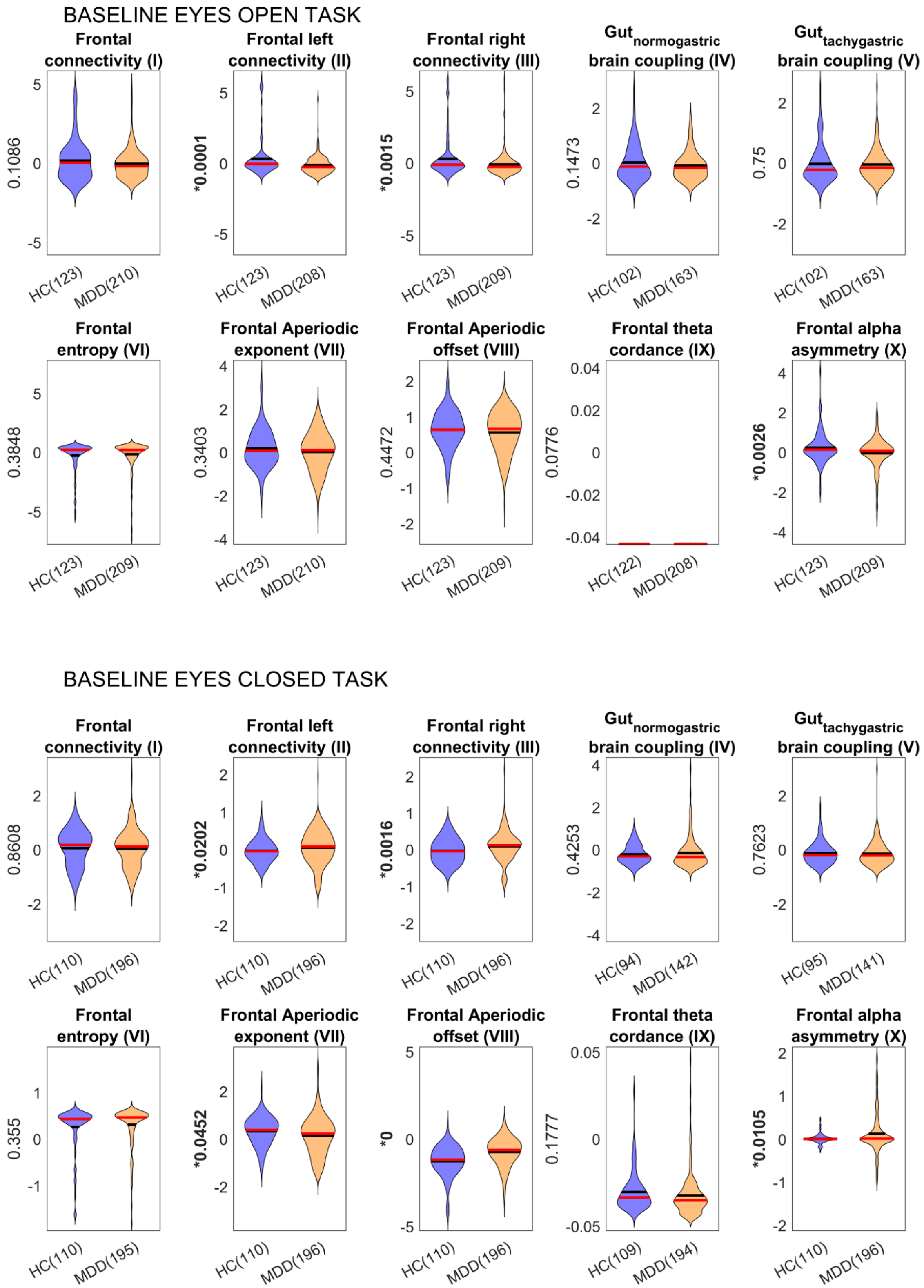

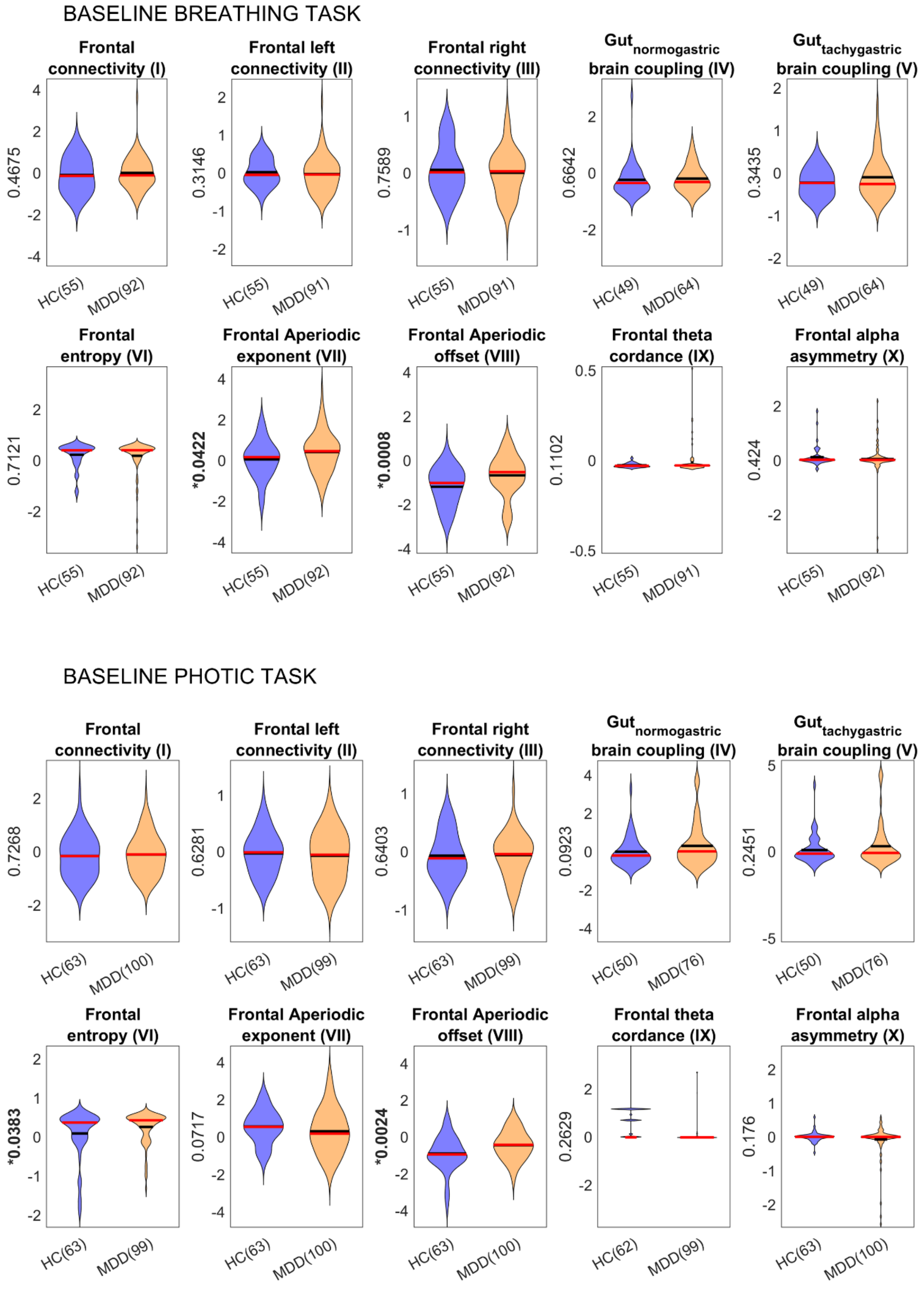


Figure II) Baseline summary feature comparison between healthy and depressed population.The p-values are uncorrected.

VISIT 2 – BASELINE EYES OPEN TASK

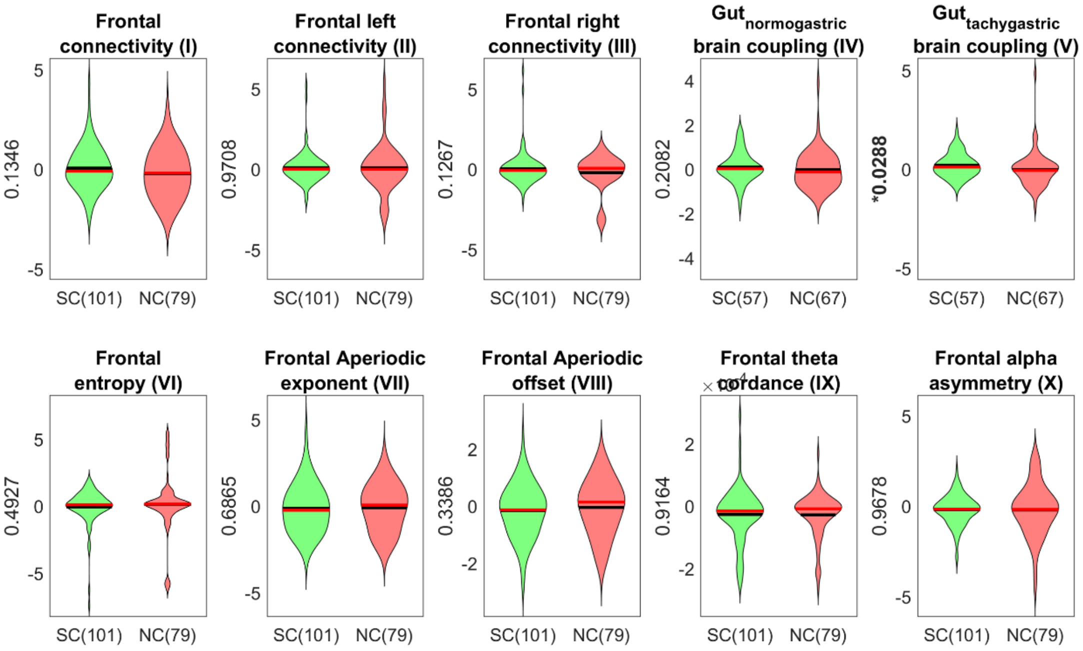


VISIT 2 - BASELINE EYES CLOSED TASK

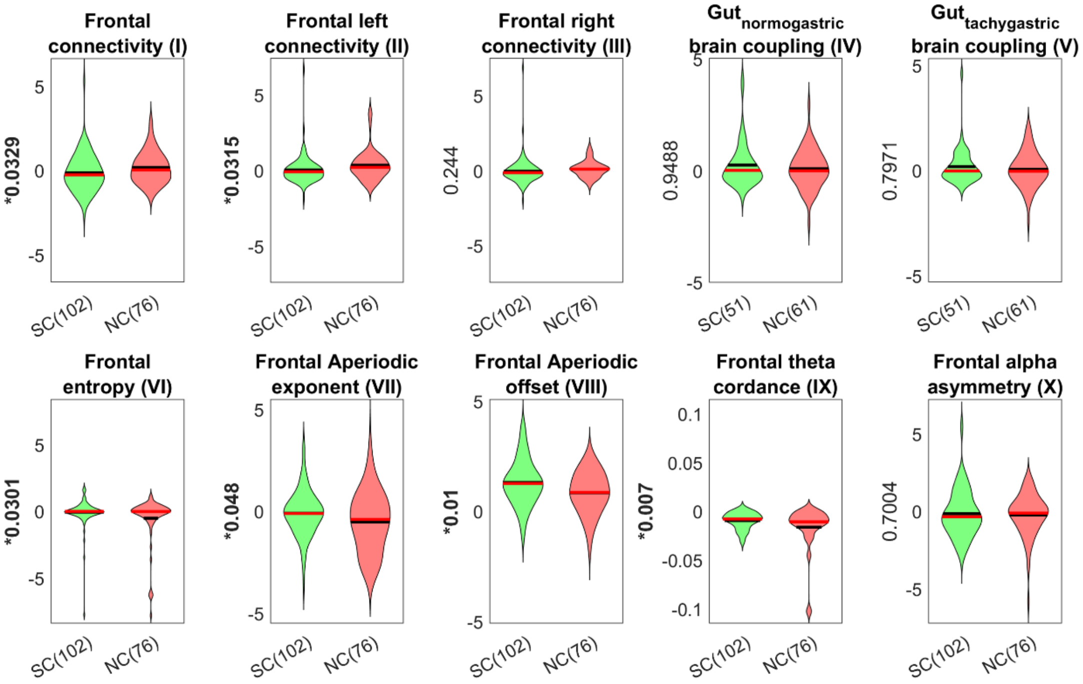

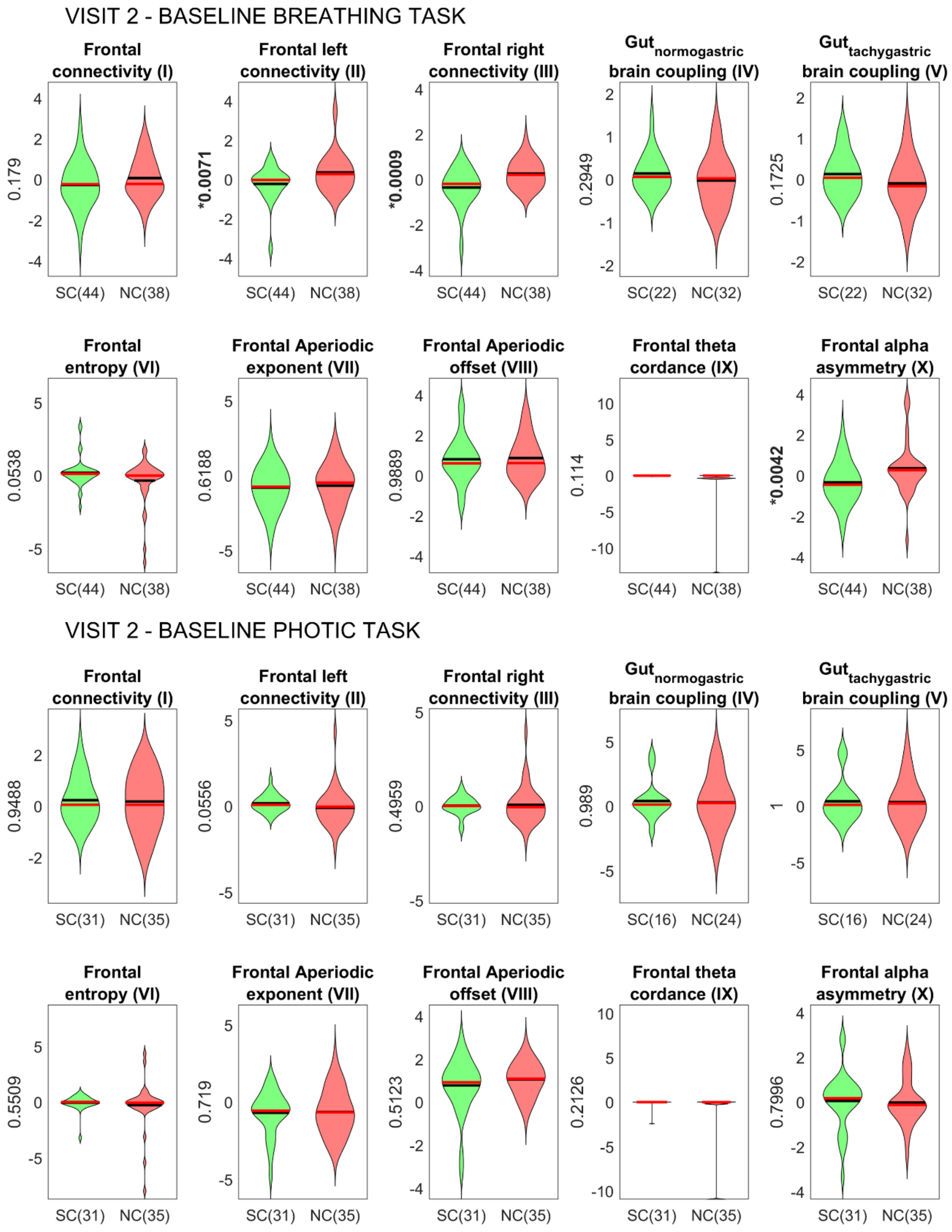


Figure III) Change in feature value at visit 2 from the baseline visit for responders and non-responders.

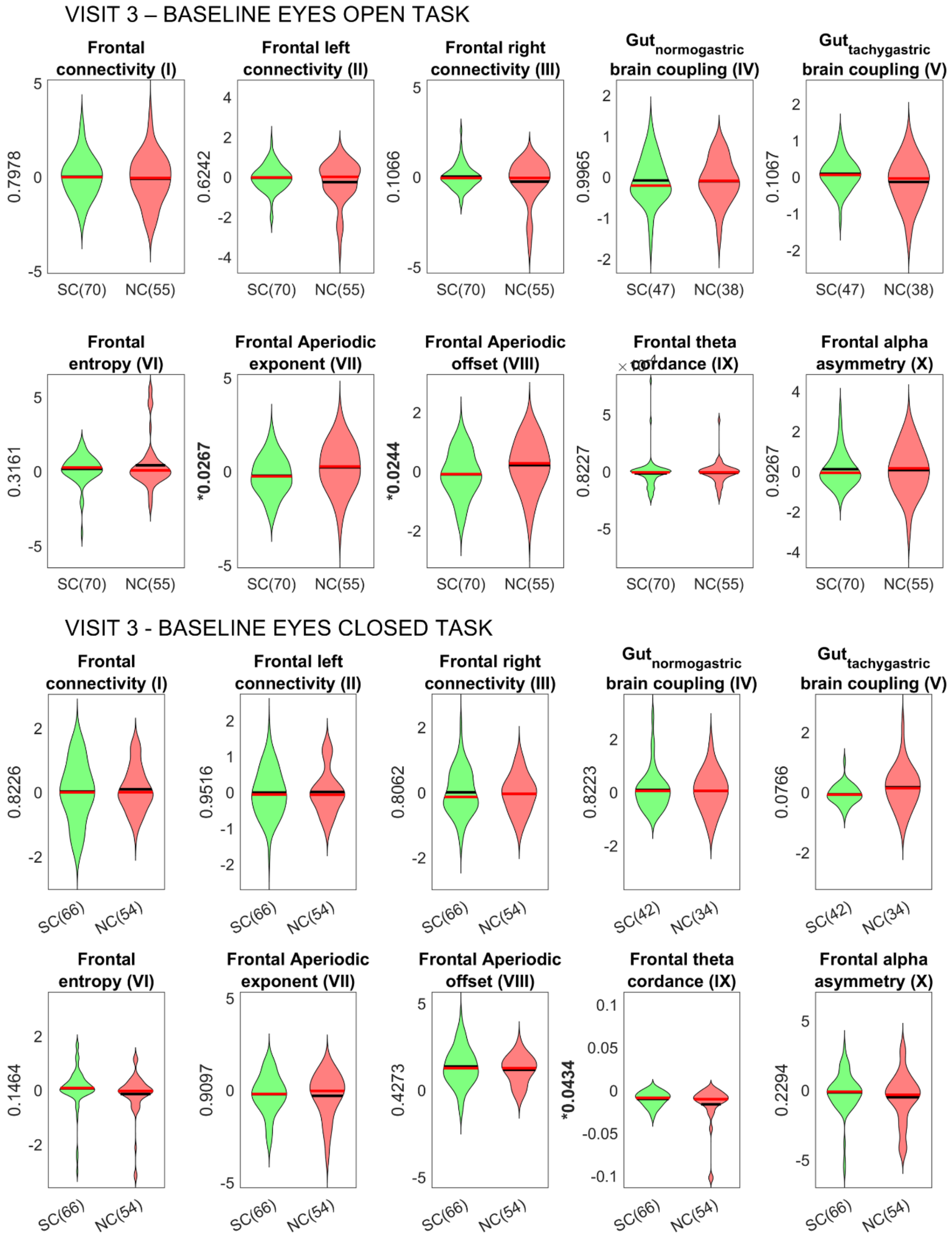
VISIT 3 – BASELINE EYES OPEN TASK
Frontal connectivity (I)
0.7978
SC(70) NC(55)
Frontal left connectivity (II)
0.6242
SC(70) NC(55)
Frontal right connectivity (III)
0.1066
SC(70) NC(55)
Gut normogastric brain coupling (IV)
0.9965
SC(47) NC(38)
Gut tachygastric brain coupling (V)
0.1067
SC(47) NC(38)
Frontal entropy (VI)
0.3161
SC(70) NC(55)
Frontal Aperiodic exponent (VII)
*0.0267
SC(70) NC(55)
Frontal Aperiodic offset (VIII)
*0.0244
SC(70) NC(55)
Frontal theta cordance (IX)
0.8227
SC(70) NC(55)
Frontal alpha asymmetry (X)
0.9267
SC(70) NC(55)
VISIT 3 - BASELINE EYES CLOSED TASK
Frontal connectivity (I)
0.8226
SC(66) NC(54)
Frontal left connectivity (II)
0.9516
SC(66) NC(54)
Frontal right connectivity (III)
0.8062
SC(66) NC(54)
Gut normogastric brain coupling (IV)
0.8223
SC(42) NC(34)
Gut tachygastric brain coupling (V)
0.0766
SC(42) NC(34)
Frontal entropy (VI)
0.1464
SC(66) NC(54)
Frontal Aperiodic exponent (VII)
0.9097
SC(66) NC(54)
Frontal Aperiodic offset (VIII)
0.4273
SC(66) NC(54)
Frontal theta cordance (IX)
*0.0434
SC(66) NC(54)
Frontal alpha asymmetry (X)
0.2294
SC(66) NC(54)

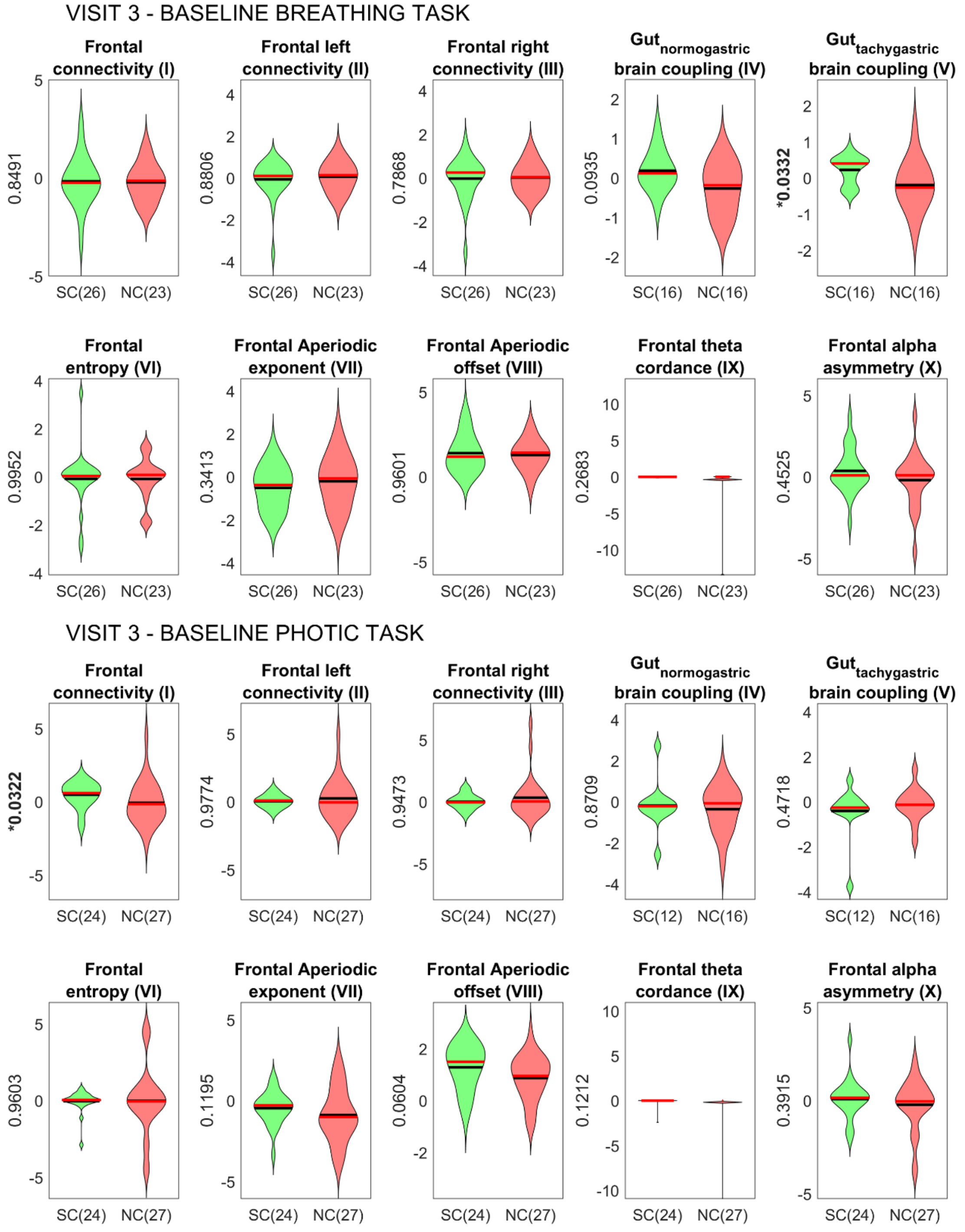


Figure IV) Change in feature value at visit 3 from the baseline visit for responders and non-responders.

From the above 3 figures (II,III,IV), we can observe that frontal connectivity with other regions, aperiodic parameters and frontal alpha asymmetry during eyes closed task are sensitive to presence of depression and also to treatment response initially. However, in the final visit, these features fail to distinguish responders and non-responders. None of the other features show reliability across more than 1 comparison for a particular task. Some of the features are distinguishing depressed population from healthy, while several other features are sensitive to treatment outcome depending on the timepoint they are probed.

#### 3) *Final list of features, groups and their importance*

| GROUP | FEATURE | SHAPLEY VALUE (NC) | SHAPLEY VALUE (SC) |
|---|---|---|---|
| - | age | 5.08E-05 | -1.16E-05 |
| - | gender | -0.00018 | -2.60E-05 |
| - | handedness | -0.0004 | 2.90E-05 |
| - | trauma | 0.001623 | 0.002374 |
| - | Patient/healthy | 0.019194 | 0.002625 |
| Social symptoms | Phq q1 v1 | 0.015715 | 0.011284 |
| Negative thoughts (self) | Phq q2 v1 | 0.000317 | 0.000627 |
| Sleep | Phq q3 v1 | 0.000407 | -0.00169 |
| Sleep | Phq q4 v1 | -0.00048 | -5.70E-05 |
| Appetite | Phq q5 v1 | -6.63E-05 | -0.00034 |
| Negative thoughts (self) | Phq q6 v1 | 0.002472 | 0.003175 |
| Social symptoms | Phq q7 v1 | 0.004395 | 0.004879 |
| Social symptoms | Phq q8 v1 | 0.000426 | -0.00057 |
| Negative thoughts (self) | Phq q9 v1 | 0.012225 | 0.009761 |
| Anxiety | Gad q1 v1 | -0.00026 | -0.00018 |
| Anxiety | Gad q2 v1 | 0.008919 | 0.006164 |

| | | | |
|---|---|---|---|
| Anxiety | Gad q3 v1 | 0.001075 | -0.00014 |
| Anxiety | Gad q4 v1 | -0.0003 | 2.63E-05 |
| Anxiety | Gad q5 v1 | 0.002266 | 0.002683 |
| Anxiety | Gad q6 v1 | 0.001378 | 0.000807 |
| Anxiety | Gad q7 v1 | 0.001451 | 0.000319 |
| Cognition | mmse category 1 | 0.000848 | 0.000434 |
| Cognition | mmse category 2 | 0.000145 | 0.000244 |
| Cognition | mmse category 3 | 2.77E-05 | 0.00072 |
| Cognition | mmse category 4 | 0.001439 | -0.00032 |
| Cognition | mmse category 5 | 0.000982 | 0.00063 |
| Cognition | mmse category 6 | -0.00037 | 0.000279 |
| Cognition | mmse category 7 | 0 | 0 |
| Cognition | mmse category 8 | -3.03E-05 | 9.72E-05 |
| - | Average of base Entropy Fl T1 base Entropy Fr T1 base Entropy Cl T1 base Entropy Cr T1 base Entropy Ol T1 base Entropy Or T1 | -4.82E-05 | 0.00052 |
| Baseline_eyes_open_global_coherence | Average of base Coh th Fl-Cl T1 base Coh th Fr-Cl T1 base Coh th Cl-Ol T1 base Coh th Cl-Or T1 base Coh alp Fl-Cl T1 base Coh alp Fr-Cl T1 base Coh alp Cl-Ol T1 base Coh bet Fl-Cl T1 base Coh bet Fr-Cl T1 base Coh bet Cl-Ol T1 base Coh bet Cl-Or T1 base Coh bet Cr-Or T1 | 0.001456 | 0.001042 |
| Baseline_eyes_open_global_coherence | Average of base Coh th Fl-Cr T1 base Coh th Fr-Cr T1 base | 0.00058 | 0.00135 |

| | | | |
|---|---|---|---|
| | Coh th Cl-Cr T1 base Coh th Cr-Ol T1 base Coh th Cr-Or T1 base Coh alp Fl-Cr T1 base Coh alp Fr-Cr T1 base Coh alp Cr-Or T1 base Coh bet Fl-Cr T1 base Coh bet Fr-Cr T1 base Coh bet Cl-Cr T1 base Coh bet Cr-Ol T1 | | |
| Baseline_eyes_open_global_coherence | Average of base Coh th Fl-Ol T1 base Coh th Fl-Or T1 base Coh th Fr-Ol T1 base Coh th Fr-Or T1 base Coh bet Fl-Ol T1 base Coh bet Fl-Or T1 base Coh bet Fr-Ol T1 base Coh bet Fr-Or T1 | -0.00338 | -0.00254 |
| Baseline_eyes_open_global_coherence | Average of base Coh th Fr-Fr T1 base Coh alp Fr-Fr T1 base Coh bet Fr-Fr T1 | -0.00183 | -0.00125 |
| Baseline_eyes_open_global_coherence | Average of base Coh th Ol-Or T1 base Coh alp Ol-Or T1 base Coh bet Ol-Or T1 | 0.008656 | 0.00677 |
| Baseline_eyes_open_global_coherence | Average of base Coh alp Fl-Ol T1 base Coh alp Fr-Ol T1 base Coh alp Fr-Or T1 | 0.000636 | 5.65E-05 |
| Baseline_Aperiodic_offset | Average of base Ap off Fl T2 base Ap off Fr T2 base Ap off Cl T2 base Ap off Cr T2 base Ap off Ol T2 base Ap off Or T2 | 0.014236 | 0.008498 |
| Baseline_relative_power | Average of base Relative Pow th Fl T2 base Relative Pow th Fr T2 base Relative Pow th Cr T2 base Relative Pow th Fl T3 base Relative Pow th Fr T3 base Relative Pow th Cl T3 base Relative Pow th Cr T3 base Relative Pow th Ol T3 base | 0.002633 | 5.33E-05 |

| | | | |
|---|---|---|---|
| | Relative Pow th Or T3 base Relative Pow th Fl T4 base Relative Pow th Fr T4 base Relative Pow th Cl T4 base Relative Pow th Ol T4 base Relative Pow th Or T4 | | |
| Baseline_relative_power | Average of base Relative Pow th Cl T2 base Relative Pow th Ol T2 base Relative Pow th Cr T4 | 0.0043 | -0.00048 |
| Baseline_relative_power | Average of base Relative Pow alp Fl T2 base Relative Pow alp Fr T2 base Relative Pow alp Cl T2 base Relative Pow alp Cr T2 base Relative Pow alp Ol T2 base Relative Pow alp Or T2 base Relative Pow alp Fl T3 base Relative Pow alp Fr T3 base Relative Pow alp Cl T3 base Relative Pow alp Cr T3 base Relative Pow alp Ol T3 base Relative Pow alp Or T3 base Relative Pow alp Fl T4 base Relative Pow alp Fr T4 base Relative Pow alp Cl T4 base Relative Pow alp Cr T4 base Relative Pow alp Ol T4 base Relative Pow alp Or T4 base alp asymmetry occipital T4 | 0.002251 | 0.00329 |
| Baseline_relative_power | Average of base Relative Pow bet Fl T2 base Relative Pow bet Fr T2 base Relative Pow bet Cl T2 base Relative Pow bet Cr T2 base Relative Pow bet Ol T2 base Relative Pow bet Or T2 base Relative Pow bet Fl T3 base Relative Pow bet Fr T3 | -0.00128 | -0.00085 |

| | | | |
|---|---|---|---|
| | base Relative Pow bet Cl T3 base Relative Pow bet Cr T3 base Relative Pow bet Ol T3 base Relative Pow bet Or T3 base Relative Pow bet Fl T4 base Relative Pow bet Fr T4 base Relative Pow bet Cl T4 base Relative Pow bet Cr T4 base Relative Pow bet Ol T4 base Relative Pow bet Or T4 | | |
| Baseline_Aperiodic_offset | Average of base Ap off Fl T3 base Ap off Fr T3 base Ap off Cl T3 base Ap off Cr T3 base Ap off Ol T3 base Ap off Or T3 | 0.005292 | 0.002566 |
| Baseline_breathing_theta_periodic_power | Average of base Periodic Pow th Cl T3 base Periodic Pow th Cr T3 base Periodic Pow th Ol T3 base Periodic Pow th Or T3 | 0.0167 | 0.003866 |
| Baseline_Aperiodic_offset | Average of base Ap off Fl T4 base Ap off Fr T4 base Ap off Cl T4 base Ap off Cr T4 base Ap off Ol T4 base Ap off Or T4 | 0.000801 | -0.00033 |
| Baseline_absolute_power | Average of base Absolute Pow th Fl T4 base Absolute Pow th Fr T4 base Absolute Pow th Cl T4 base Absolute Pow th Cr T4 base Absolute Pow th Ol T4 base Absolute Pow th Or T4 base Absolute Pow alp Fl T4 base Absolute Pow alp Fr T4 base Absolute Pow alp Cr T4 base Absolute Pow alp Ol T4 base Absolute Pow alp Or T4 base Absolute Pow bet Fl T4 base Absolute Pow bet Fr T4 base Absolute Pow bet Cl T4 base Absolute Pow bet Cr T4 | 0.000671 | 0.000101 |

| | | | |
|---|---|---|---|
| | base Absolute Pow bet Ol T4 base Absolute Pow bet Or T4 | | |
| Baseline_eyes_open_global_coherence | base Coh th Cr-Cr T1 | 0.004501 | 0.001367 |
| Baseline_eyes_open_global_coherence | base Coh alp Fl-Fr T1 | 0.003714 | 0.002794 |
| Baseline_eyes_open_global_coherence | base Coh alp Cl-Cr T1 | -0.00044 | -0.00048 |
| Baseline_eyes_open_global_coherence | base Coh alp Cl-Or T1 | 0.016104 | 0.012517 |
| Baseline_eyes_open_global_coherence | base Coh alp Cr-Ol T1 | -0.00167 | -0.00089 |
| - | base Entropy Ol T2 | -0.00046 | 0.000148 |
| Baseline_eyes_closed_central_left_coherence | base Coh th Cl-Or T2 | 0.003177 | -0.00067 |
| Baseline_eyes_closed_non_central_left_coherence | base Coh bet Fl-Fl T2 | 0.002368 | -0.00271 |
| - | base PLI Fl-Fr T2 | 0.005308 | 0.003767 |
| - | base PLI Ol-Ol T2 | 0.003825 | 0.005516 |
| Baseline_breathing_low_frequency_coherence | base Coh alp Fl-Cl T3 | 0.014864 | 0.013338 |
| Baseline_breathing_low_frequency_coherence | base Coh alp Fr-Cr T3 | 0.003845 | 0.003262 |
| Baseline_breathing_low_frequency_coherence | base Coh alp Cr-Ol T3 | -0.0002 | -0.00061 |
| Baseline_breathing_central_tachy_PAC | base tachy PAC bet top Cr T3 | 0.004111 | 0.001112 |
| Baseline_photic_central_left_coherence | base Coh th Fr-Cl T4 | 0.000875 | 0.001967 |

| | | | |
|---|---|---|---|
| Baseline_photic_non_central_left_coherence | base Coh alp Fr-Fr T4 | -0.00098 | -3.34E-05 |
| Baseline_photic_non_central_left_coherence | base Coh alp Fr-Cr T4 | 0.004286 | 0.001904 |
| Baseline_photic_non_central_left_coherence | base Coh alp Cr-Ol T4 | 0.009384 | 0.002897 |
| Baseline_photic_non_central_left_coherence | base Coh bet Or-Or T4 | 0.003627 | -0.00329 |
| Longitudinal_Aperiodic_offset | Average of Ap exp Fl T1 Ap off Fl T1 | -0.00082 | -0.00091 |
| Longitudinal_Aperiodic_offset | Average of Ap off Ol T1 Ap off Or T1 | -0.00061 | -0.00093 |
| Longitudinal_eyes_open_fronto-central_left_beta_periodic_power | Average of Periodic Pow bet Fl T1 Periodic Pow bet Cl T1 | 0.002073 | 0.001369 |
| - | Average of Periodic Pow bet Cr T1 Periodic Pow bet Ol T1 Periodic Pow bet Or T1 | 0.003686 | 0.003711 |
| Longitudinal_fronto-central_left_absolute_power | Average of Absolute Pow th Cl T1 th cordance Cl T1 | 0.00011 | 6.06E-05 |
| Longitudinal_eyes_open_global_coherence | Average of Coh th Cr-Ol T1 Coh bet Cr-Ol T1 | 0.003978 | 0.001764 |
| Longitudinal_eyes_open_global_coherence | Average of Coh th Ol-Or T1 Coh alp Ol-Or T1 Coh bet Ol-Or T1 | -0.0003 | 0.001782 |
| - | Average of Relative Pow bet Fl T2 Relative Pow bet Fr T2 Relative Pow bet Cl T2 Relative Pow bet Cr T2 Relative Pow bet Ol T2 Relative Pow bet Or T2 Relative Pow bet Fr T3 Relative Pow bet Cl T3 Relative Pow bet | 0.000907 | -0.00232 |

| | | | |
|---|---|---|---|
| | Cr T3 Relative Pow bet Ol T3 Relative Pow bet Or T3 | | |
| Longitudinal_eyes_closed_central_left_coherence | Average of Coh th Cl-Cl T2 Coh bet Cl-Cl T2 | 0.004102 | -0.00147 |
| Longitudinal_fronto-central_left_absolute_power | Average of Absolute Pow th Fl T3 Absolute Pow alp Fl T3 Absolute Pow alp Cl T3 Absolute Pow alp Cr T3 Absolute Pow alp Ol T3 Absolute Pow alp Or T3 Absolute Pow bet Fl T3 Absolute Pow bet Fr T3 Absolute Pow bet Cl T3 Absolute Pow bet Cr T3 | 0.004003 | 0.003358 |
| Longitudinal_photic_non_central_left_coherence | Average of Coh th Fl-Fl T4 Coh bet Fl-Fl T4 | 0.006108 | 0.010807 |
| Longitudinal_photic_non_central_left_coherence | Average of Coh th Fl-Cr T4 Coh th Cr-Or T4 | 0.005073 | 0.003881 |
| Longitudinal_Aperiodic_offset | Ap exp Ol T1 | 0.010496 | 0.004676 |
| Longitudinal_eyes_open_fronto_left_beta_relative_power | Relative Pow bet Fl T1 | 0.001331 | -0.00235 |
| Longitudinal_eyes_open_global_coherence | Coh bet Cr-Cr T1 | 0.004614 | 0.001228 |
| - | th asymmetry occipital T1 | -0.0014 | -0.00107 |
| Longitudinal_eyes_closed_central_left_coherence | Coh th Cl-Ol T2 | 0.010509 | 0.002536 |
| Longitudinal_eyes_closed_non_central_left_coherence | Coh th Cr-Cr T2 | 0.002934 | 0.000754 |
| Longitudinal_eyes_closed_non_central_left_coherence | Coh alp Fl-Fr T2 | 0.009293 | 0.006183 |
| Longitudinal_eyes_closed_non_central_left_coherence | Coh alp Fl-Ol T2 | 0.002592 | 0.004229 |

| | | | |
|---|---|---|---|
| Longitudinal_eyes_closed_non_central_left_coherence | Coh alp Fl-Or T2 | 0.001896 | 0.001337 |
| Longitudinal_eyes_closed_non_central_left_coherence | Coh alp Fr-Or T2 | 0.016035 | 0.012844 |
| Longitudinal_eyes_closed_central_left_coherence | Coh alp Cl-Cl T2 | 0.003974 | -0.00133 |
| Longitudinal_eyes_closed_non_central_left_coherence | Coh alp Or-Or T2 | 0.004425 | -0.00252 |
| Longitudinal_eyes_closed_central_left_coherence | Coh bet Cl-Ol T2 | 0.008804 | -0.00046 |
| Longitudinal_eyes_closed_non_central_left_coherence | Coh bet Cr-Cr T2 | 0.003715 | -6.80E-06 |
| Longitudinal_eyes_closed_non_central_left_coherence | Coh bet Cr-Ol T2 | 0.001588 | -0.00275 |
| - | PLI Fl-Or T2 | 0.002502 | 0.000891 |
| - | PLI Fr-Fr T2 | 0.000541 | -0.00061 |
| Longitudinal_normo_PAC | normo PAC alp top Fl T2 | 0.002504 | 0.001151 |
| Longitudinal_normo_PAC | normo PAC alp top Ol T2 | 0.000525 | -0.0009 |
| - | tachy PAC th top Fl T2 | -4.27E-05 | 0.001114 |
| - | tachy PAC alp top Fl T2 | 0.006698 | 0.006527 |
| - | th asymmetry occipital T2 | 0.004749 | 0.001275 |
| Longitudinal_low_frequency_central_asymmetry | alp asymmetry central T2 | 0.023277 | 0.019103 |
| Longitudinal_high_frequency_frontal_asymmetry | beta asymmetry frontal T2 | 0.022208 | 0.009193 |
| Longitudinal_breathing_fronto-central_left_theta_periodic_power | Periodic Pow th Cl T3 | 0.04386 | 0.029372 |

| | | | |
|---|---|---|---|
| Longitudinal_breathing_fronto_left_beta_relative_power | Relative Pow bet Fl T3 | 0.003048 | -0.00226 |
| Longitudinal_breathing_low_frequency_coherence | Coh th Cr-Cr T3 | 0.000249 | -0.00165 |
| Longitudinal_breathing_low_frequency_coherence | Coh alp Fl-Cr T3 | -0.00456 | 0.000362 |
| Longitudinal_breathing_high_frequency_coherence | Coh bet Fr-Cr T3 | 0.001184 | -0.00212 |
| Longitudinal_breathing_high_frequency_coherence | Coh bet Fr-Ol T3 | 0.017992 | 0.012467 |
| Longitudinal_normo_PAC | normo PAC brd top Or T3 | 0.001228 | 0.001127 |
| Longitudinal_normo_PAC | normo PAC alp top Or T3 | 0.003027 | 0.006878 |
| - | tachy PAC alp top Or T3 | 0.013614 | 0.012587 |
| Longitudinal_breathing_central_tachy_PAC | tachy PAC bet top Cl T3 | 0.001239 | 0.000692 |
| Longitudinal_breathing_central_tachy_PAC | tachy PAC bet top Cr T3 | 0.00763 | 0.005899 |
| - | tachy PAC bet top Or T3 | -0.00103 | -0.00022 |
| Longitudinal_high_frequency_frontal_asymmetry | beta asymmetry frontal T3 | 0.028463 | 0.011418 |
| Longitudinal_photic_central_left_coherence | Coh th Cl-Cr T4 | 0.001768 | 2.00E-05 |
| Longitudinal_photic_central_left_coherence | Coh alp Cl-Or T4 | 0.007817 | 0.004112 |
| Longitudinal_photic_non_central_left_coherence | Coh alp Ol-Or T4 | 0.007304 | 0.002176 |
| Longitudinal_photic_central_left_coherence | Coh bet Fl-Cl T4 | 0.00318 | 0.003703 |

| | | | |
|---|---|---|---|
| Longitudinal_photic_central_left_coherence | Coh bet Fr-Cl T4 | 0.00918 | 0.007004 |
| Longitudinal_photic_central_left_coherence | Coh bet Cl-Ol T4 | 0.000331 | -0.00228 |
| Longitudinal_photic_non_central_left_coherence | Coh bet Ol-Or T4 | 0.003007 | 0.001013 |
| - | PLI Cl-Cl T4 | -0.00325 | 2.08E-05 |
| - | PLI Cr-Ol T4 | -0.00039 | 0.000568 |
| Longitudinal_low_frequency_central_asymmetry | th asymmetry central T4 | 0.008169 | 0.000334 |

Table A: List of selected features and their groups for feature importance. T1- eyes open, T2- eyes closed, T3- breathing, T4- photic administration tasks.

*Feature importance*

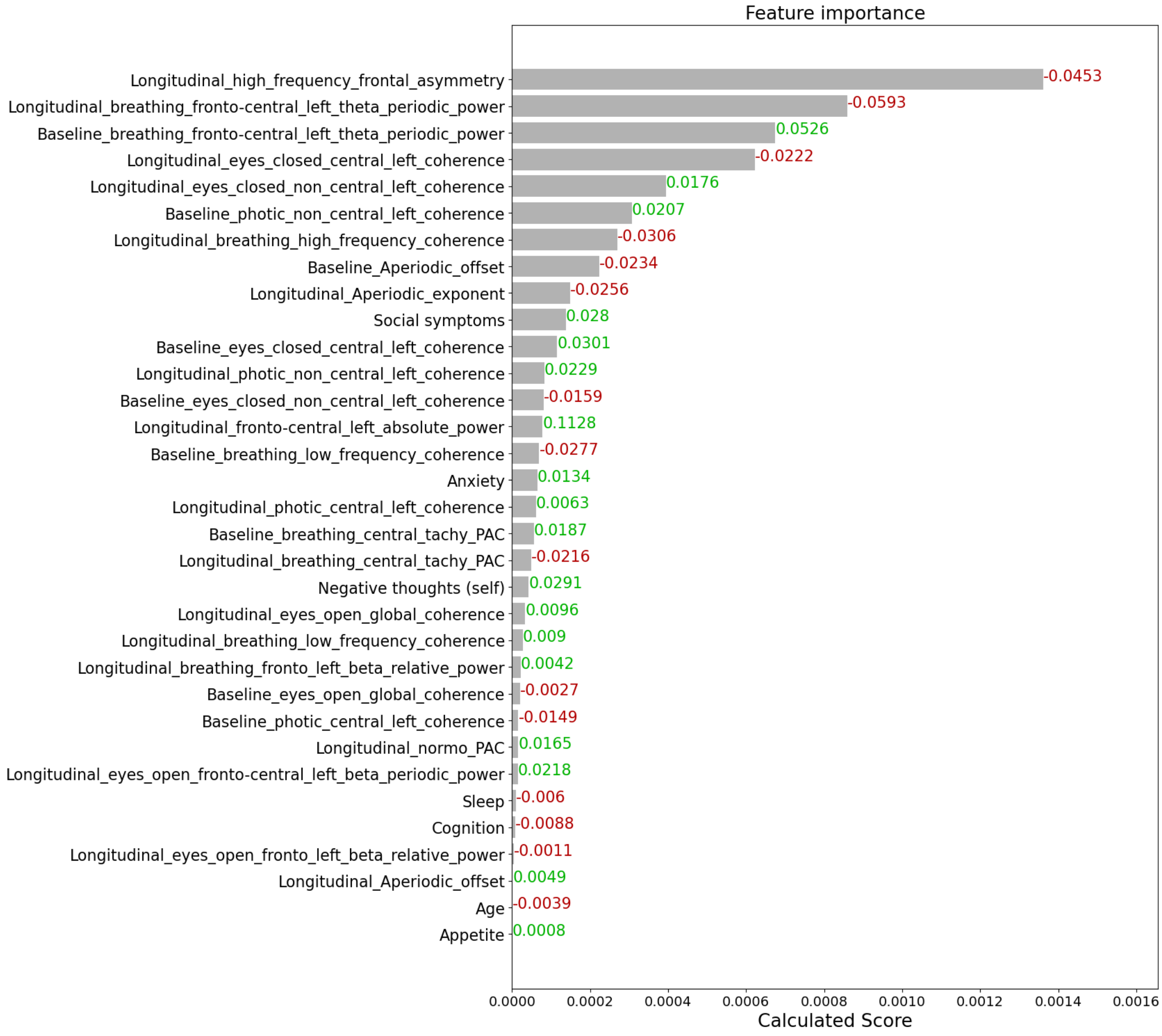


Figure V: Feature importance based on calculated score and grouping.

The slope depicts the effect of changing the feature value on shapley value. Red color depicts that increasing the feature value decreases the marginal contribution towards response and vice versa for green.

#### *4) Symptom wise electrophysiological feature*

Significant reduction in coupling of the tachygastric gut rhythm with the beta signals of central brain region, and also in overall beta coherence during breathing task, were seen in responders who had manifested higher severity than population median in appetite symptoms, negative thought about self, or social symptoms. Similarly, frontal beta asymmetry and periodic theta power in left fronto-central regions while breathing task showed significant reduction in responders with relatively higher appetite, sleep and social issues in the population. Responders with sleep issues displayed significant increase in coherence during photic administration task. Interestingly, responders with anxiety showed significant decrease in coherence in central left region, while it increased in the other regions during resting state eyes-closed task. Overall, we observed different electrophysiological biomarkers to be predictive of treatment outcome depending on their psychotype and physiological symptom presentation (**Table B**).

| **Symptom group** | **Electrophysiology feature** | **mean_NC** | **mean_SC** | **corrected p_value** |
|---|---|---|---|---|
| Appetite | Baseline breathing left fronto-central theta periodic power | -0.160 (± 0.432) | 0.700 (± 1.146) | 0.034 |
| | Baseline photic left non central coherence | -0.542 (± 0.294) | -0.104 (± 0.365) | <0.001 |
| | Longitudinal breathing central tachy PAC | 0.282 (± 0.596) | -0.176 (± 0.379) | 0.042 |
| | Longitudinal breathing beta coherence | 0.192 (± 0.115) | -0.278 (± 0.328) | <0.001 |
| | Longitudinal beta frontal asymmetry | 0.259 (± 0.712) | -0.612 (± 1.121) | 0.033 |
| | Longitudinal breathing left fronto-central theta periodic power | 0.235 (± 0.715) | -0.969 (± 0.823) | <0.001 |
| Anxiety | Longitudinal eyes closed left non central coherence | -0.323 (± 0.330) | 0.377 (± 0.776) | <0.001 |
| | Longitudinal eyes closed left central coherence | 0.261 (± 0.291) | 0.063 (± 1.238) | 0.041 |
| Negative thoughts self | Longitudinal breathing central tachy PAC | 0.356 (± 0.611) | -0.143 (± 0.395) | 0.020 |
| | Longitudinal eyes closed left central coherence | 1.154 (± 1.946) | 0.032 (± 1.259) | 0.031 |
| | Longitudinal breathing beta coherence | 0.772 (± 0.793) | -0.054 (± 0.469) | <0.001 |
| Sleep | Longitudinal photic non central beta coherence | -0.196 (± 0.461) | 0.222 (± 0.449) | 0.003 |

| | | | | |
|---|---|---|---|---|
| | Longitudinal photic central beta coherence | -0.070 (± 0.299) | 0.248 (± 0.525) | 0.040 |
| | Longitudinal beta frontal asymmetry | 0.105 (± 0.833) | -0.588 (± 1.037) | 0.006 |
| | Longitudinal breathing left fronto-central theta periodic power | 0.078 (± 0.753) | -0.825 (± 0.748) | <0.001 |
| Social symptoms | Longitudinal breathing central tachy PAC | 0.291 (± 0.565) | -0.130 (± 0.390) | 0.039 |
| | Longitudinal eyes closed left central coherence | 0.723 (± 1.763) | -0.106 (± 1.150) | 0.024 |
| | Longitudinal breathing beta frequency coherence | 0.501 (± 0.559) | -0.039 (± 0.414) | <0.001 |
| | Longitudinal beta frontal asymmetry | 0.315 (± 0.512) | -0.480 (± 0.790) | <0.001 |
| | Longitudinal breathing left fronto central theta periodic power | 0.370 (± 0.910) | -0.753 (± 0.505) | <0.001 |

**Table B: Significant features for specific phenotypic subtypes-** Electrophysiological feature comparison for responders and non-responders in a subset of population with specific symptom severity, with relatively higher symptom score than the population median. The above table highlights the feature group from the top 20 that are significant after Bonferonni correction for multiple comparisons, their corrected p-value, the mean feature value and N of SC and NC groups exhibiting significant change and no change in mental health for the specific subset of cohort.
